# A schedule-driven importation risk framework for mass gathering events: FIFA World Cup 2026

**DOI:** 10.64898/2026.06.03.26354828

**Authors:** Jose L. Herrera-Diestra, Nyall Jamieson, Mallory J. Harris, Kaiming Bi, Martial L. Ndeffo-Mbah, Melissa Zeynep Ertem, Amera Al-Amery, Lauren Ancel Meyers

## Abstract

**Objectives:** Large international events draw visitors worldwide, but how much do they add to infectious disease importation risk beyond everyday travel? We developed a schedule-driven, uncertainty-aware framework to estimate this added risk and applied it to the 2026 FIFA World Cup as a case study.

**Methods:** Three nested Poisson models represent baseline travel, event-related growth, and schedule-driven allocation of additional visitors by venue and date, with Monte Carlo simulation propagating uncertainty in disease under-reporting and travel-while-infected probability. We applied the approach to five diseases across 11 US host cities during the June 2026 World Cup group stage using only publicly available data, evaluated against independently reported dengue case counts, with measles as an additional qualitative check.

**Results:** Relative to 2026 travel without the tournament, the World Cup added 1.0–6.5% to expected importations under schedule-driven routing, versus 2.9–8.5% under routing that ignores the match schedule. Dengue, malaria, and pertussis had the highest modeled importation levels, and the probability of at least one infected dengue or pertussis arrival exceeded 0.99 at 10 of 11 cities. Comparison with 2024 dengue and 2022 measles surveillance data showed a significant rank correlation between modeled and reported dengue cases (Spearman *ρ* = 0.83).

**Conclusions:** This approach provides an empirically grounded way to assess importation risk during large international events using only freely available data, identifying where public health planning may be most useful. For the World Cup case study, the tournament’s own contribution was modest once fans were routed by the actual match schedule rather than as ordinary travelers.

## 1 Introduction

Mass gathering events concentrate infectious disease importation risk by drawing large, diverse international populations into a small number of cities over a short period. Quantifying that risk in a way that is reusable across events, transparent about uncertainty, and buildable from routinely collected data, rather than through a bespoke, one-off analysis, remains an open methodological need. We address this need with a schedule-driven importation risk framework, and demonstrate it using the 2026 FIFA World Cup (June 11 – July 19, 2026) as a case study, the first edition to include 48 participating nations, played across 16 venues in the United States, Canada, and Mexico. The 11 US host cities are Atlanta, Boston, Dallas, Houston, Kansas City, Los Angeles, Miami, New York, Philadelphia, San Francisco, and Seattle. The geographic distribution of all 16 WC2026 stadiums across the three host nations is shown in Supplementary Figure S4.

Importation risk analyses preceding past FIFA World Cups have taken different approaches depending on the host country’s disease ecology. Ahead of the 2014 tournament in Brazil, van Panhuis et al. estimated the number of dengue infections that international visitors would acquire in host cities, using host-city incidence rates and ticket allocations, and reported uncertainty ranges for these estimates [1]. That analysis addressed infection risk for visitors to an endemic host country; the present framework addresses the reverse direction, the importation of infections by visitors into host cities. For Russia 2018, Pshenichnaya et al. reviewed infection risks for attendees, including gastrointestinal infections, tick-borne encephalitis and Lyme disease, sexually transmitted infections, measles, and rarer risks such as rabies and West Nile fever, while noting that large sporting events have rarely been associated with infectious disease outbreaks [2]. For Qatar 2022, Al-Tawfiq et al. reviewed infection risks associated with the tournament, including COVID-19, mpox, MERS coronavirus, measles, and, at low incidence, dengue [3]. A concurrent multi-pathogen assessment for WC2026 by Lessler et al. [4] evaluated 77 pathogens and found the World Cup’s added risk to be near zero for 63 of them, with the remaining priority pathogens falling into five categories that include mosquito-borne pathogens and seasonal respiratory viruses. The present study is complementary to that breadth-first screening; rather than covering the full pathogen space, we provided city-specific quantitative importation estimates with propagated parameter uncertainty for five diseases. The current analysis focuses on the group-stage period (June 2026) rather than the full tournament window extending into July, as travel volumes and disease incidence inputs are calibrated to June.

Two methodological gaps motivated this study. First, country-level air travel routing data from the Bureau of Transportation Statistics T-100 program enable city-specific risk stratification at all 11 US venues. Second, literature ranges for under-reporting correction factors and travel-while-infected probabilities span several-fold; propagating that uncertainty into estimates is essential for honest risk communication.

We address these gaps with a three-tier nested Poisson importation model with 5,000draw Monte Carlo uncertainty for five diseases (dengue, malaria, measles, pertussis, influenza). Every input (air-travel routing via BTS T-100, border arrivals via CBP I-94, travel growth forecasts via NTTO, disease incidence via WHO, and parameter ranges from published studies and CDC references) is free and publicly available, with no proprietary mobility datasets, commercial travel-intelligence subscriptions, or specialized simulation infrastructure required. The approach is therefore designed to be directly usable by public health agencies, including those without dedicated modeling staff or budget for commercial data, as a rapidly deployable first-pass risk assessment for any future mass gathering event, complementing more resource-intensive frameworks where those resources are available.

## 2 Methods

The framework quantifies importation risk for a set of diseases at a set of host locations during a defined event period, using three nested Poisson models that share identical disease data so that differences in results reflect travel-volume assumptions only. We first describe the framework in general terms, then its application to the case study of five infectious diseases (dengue, malaria, measles, pertussis, and influenza) at the 11 US FIFA World Cup 2026 host cities during June 2026.

### 2.1 General framework

For a host location *v* receiving visitors from source location *c* during an event period, three nested travel-volume tiers are distinguished.

**Baseline (M1)** projects arrivals from ordinary background travel growth over the period leading up to the event, with no event-specific increment, representing travel volumes in the event’s absence.

**Event-adjusted (M2)** applies an aggregate event-driven growth factor *ϕ_c_* to project arrivals, distributed across host locations by the same background routing used in M1; this represents the total scale of event-attributable travel without allocating it to the specific host locations where event attendees actually go.

**Schedule-driven (M3)** decomposes projected arrivals into the background stream (routed as in M1) and an event-specific visitor stream, allocated to host locations by an eventschedule-driven routing fraction *ω_c,v_* that reflects where and when source-location *c*’s attendees are expected to arrive, rather than by ordinary background routing. Because an event’s own schedule (match venues, festival stages, pilgrimage sites) typically determines where its visitors go, M3 is intended to give a more realistic host-location risk distribution, redirecting the event-specific stream away from host locations where the event does not occur.

Comparing M1 to M3 isolates the event-specific increment under realistic, schedule-driven allocation, cleanly, because both use the same projected future travel scale rather than an unprojected historical baseline. Comparing M2 to M3 isolates the effect of the allocation choice itself, since M2 shows what the same event-driven growth would imply if routed like ordinary background travel instead of by the event’s actual schedule, an estimate that misallocates the event’s contribution among host locations and can also misstate its total across them (Appendix A.7). Full mathematical derivations of the projected number of arrivals *N_c,v_* under each tier, and the general definition of the routing fractions, are provided in Appendix A of the Supplementary Material.

For each source location *c*, host location *v*, and disease *d*, the expected number of infected arrivals is:

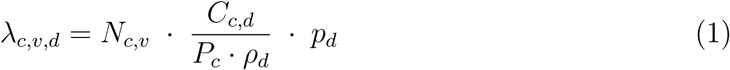

where *N_c,v_* is the projected number of arrivals under a given model tier, *C_c,d_/P_c_* is the reported per-capita incidence, *ρ_d_* is the source-location reporting fraction (the estimated proportion of true infections captured by surveillance, so dividing by *ρ_d_* corrects reported incidence up to true incidence), and *p_d_* is the probability that a person infected during the incidence period is still infected, and not yet too ill to travel, at the time of travel (defined in Section 2.2). Host-location intensity and importation probability follow as Λ*_v,d_* = Σ*_c_λ_c,v,d_* and *P* (*≥* 1)*_v,d_* = 1 *−* exp(*−*Λ*_v,d_*), where *X_v,d_ ∼* Poisson(Λ*_v,d_*) is the number of imported cases at location *v* for disease *d*; for brevity we write this as *P* (*≥* 1)*_v,d_*. Λ*_v,d_* is an expected number of infected arrivals, detected or not, and not a prediction of diagnosed US cases (see Discussion). This formulation, and the three model tiers above, apply to any event, disease set, and set of host locations for which travel volumes, disease incidence, and an event-schedule allocation rule are available (see Discussion, Generalizability).

### 2.2 Application to the 2026 FIFA World Cup

We applied this framework to five infectious diseases (dengue, malaria, measles, pertussis, and influenza) at each of the 11 US FIFA World Cup 2026 host cities during June 2026, the tournament’s group-stage period. For this application, source and host locations are countries of origin and US host cities respectively, and the general Event-adjusted tier (M2) is instantiated as a World Cup-adjusted model, referred to as such throughout the remainder of the paper.

#### Travel data

Country-of-residence (COR) monthly arrivals were obtained from the US Customs and Border Protection I-94 program [5]. City-level routing fractions were derived from BTS T-100 International Segment (Form 41 Traffic) nonstop passenger records for June 2023–2025 [6], computed as the share of each country’s US-bound passengers that landed at each venue city, pooled over the three Junes for stability; these fractions provide the background routing used by M1 and M2. National Travel and Tourism Office (NTTO) projections provided the event-driven growth factor *ϕ_c_* for the 12 largest US inbound source markets [7]; a global World Cup factor of 1.174 was applied to other qualified nations, and an empirical background trend factor to non-qualified countries. The event-schedule routing fraction *ω_c,v_* (M3) followed each qualified team’s actual group-stage match venues, redirecting the fan stream to Canadian or Mexican venues when a team played there rather than distributing it across all 11 US cities (Appendix A.4).

#### Disease data

Disease-specific incidence was sourced from the WHO Global Dengue Surveillance dataset (monthly; June 2024–2025 average) [8], the WHO World Malaria Report 2024 (annual incidence per 1,000 population) [9], WHO Immunization Data (annual incidence per million for measles and pertussis) [10], and WHO FluNet GISRS (weekly positive specimens, ISO weeks 22–26, 2023–2025 average) [11]. For influenza, *C_c,d_* is therefore the number of laboratory-confirmed positive specimens reported to FluNet, which depends on each country’s testing volume as well as on influenza activity, so *ρ_d_* acts as a ratio of positive specimens to infections and influenza estimates are more uncertain than the range in Table 1 implies. National population estimates *P_c_* used 2026 projections [12]. Literature-derived disease parameters are summarized in Table 1.

**Table 1:**
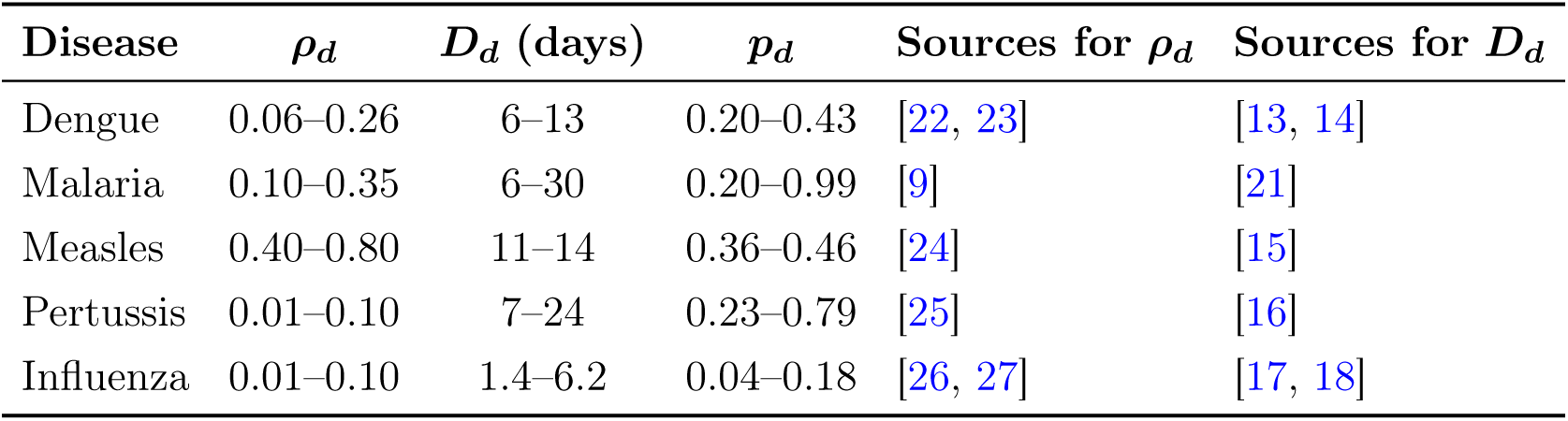
Disease parameters and Monte Carlo uncertainty bounds used in all three model tiers. The travel-while-infected probability is *p_d_* = *D_d_/T_d_*, where *D_d_* is the travel-eligible window in days, bounded below by the incubation period and above by the incubation period plus the subsequent period compatible with travel (for malaria, the incubation period only; see Methods), and *T_d_* is the length of the June incidence period (30.4 days; 35 days for influenza, whose FluNet input covers ISO weeks 22–26). For each of 5,000 iterations, *ρ_d_ ∼* Uniform(*ρ*_min_*, ρ*_max_) and *D_d_ ∼* Uniform(*D*_min_*, D*_max_) are drawn independently. All reported estimates are Monte Carlo medians with 95% uncertainty intervals. Dengue incidence and the influenza FluNet input correspond to the June analysis period; malaria, measles, and pertussis use annual rates divided by 12 to approximate June incidence.

| Disease | $\rho_d$ | $D_d$ (days) | $p_d$ | Sources for $\rho_d$ | Sources for $D_d$ |
| --- | --- | --- | --- | --- | --- |
| Dengue | 0.06–0.26 | 6–13 | 0.20–0.43 | [22, 23] | [13, 14] |
| Malaria | 0.10–0.35 | 6–30 | 0.20–0.99 | [9] | [21] |
| Measles | 0.40–0.80 | 11–14 | 0.36–0.46 | [24] | [15] |
| Pertussis | 0.01–0.10 | 7–24 | 0.23–0.79 | [25] | [16] |
| Influenza | 0.01–0.10 | 1.4–6.2 | 0.04–0.18 | [26, 27] | [17, 18] |

#### Travel-while-infected probability

We write *p_d_* = *D_d_/T_d_*, where *D_d_* is the mean number of days an infection remains travel-eligible, from infection until recovery or until illness prevents travel, and *T_d_* is the length of the period over which *C_c,d_* is counted (30.4 days for June, and 35 days for influenza, whose FluNet input covers ISO weeks 22–26). The fraction of infections that stay mild enough to travel through is rarely measured, so we bounded *D_d_* with two well-documented quantities (Table 1). The lower bound is the incubation period, because any infected person can travel while incubating. The upper bound adds the subsequent period during which infection typically remains compatible with travel, namely viremia for dengue, the prodrome before rash onset for measles, the catarrhal stage for pertussis, and viral shedding for influenza. This gives 6–13 days for dengue (incubation mean 5.9 days, 95% within 3–10 days, and viremia of about 7 days) [13, 14], 11–14 days for measles (11–12 days from exposure to prodrome and about 14 days to rash onset) [15], 7–24 days for pertussis (incubation commonly 7–10 days and a catarrhal stage of 1–2 weeks) [16], and 1.4–6.2 days for influenza (median incubation of 1.4 days for influenza A and viral shedding of 4.8 days) [17, 18]. For malaria, untreated *Plasmodium falciparum* infections in endemic settings can persist asymptomatically for hundreds of days [19, 20], so we restricted the window to the usual incubation period of 6–30 days [21], counting infections that can become clinical after arrival and excluding long-term asymptomatic carriage.

### 2.3 Monte Carlo uncertainty quantification

The under-reporting correction *ρ_d_* and the travel-eligible window *D_d_* each have wide ranges, reflecting heterogeneity across surveillance systems and clinical courses. We treated both as independent Uniform random variables within their bounds (Table 1) and drew 5,000 samples per disease. The same 5,000 draws were used for all three model tiers of a given disease, so that differences between tiers reflect travel-volume assumptions only and are not affected by Monte Carlo sampling noise. For each draw, all city-level Λ*_v,d_* values were recomputed. Reported estimates are Monte Carlo medians with 95% uncertainty intervals (UIs; 2.5th–97.5th percentiles). Because *ρ_d_* and *p_d_* enter multiplicatively, uncertainty intervals are asymmetric, and the direction of the asymmetry is disease-specific rather than uniform. Dengue, measles, pertussis, and influenza show wider upper intervals (right-skewed, with uncertainty extending farther above the median than below it), whereas malaria shows wider lower intervals (left-skewed, with uncertainty extending farther below the median) (Supplementary Figure S3).

Because Poisson arrivals are assumed independent, correlated group travel could inflate the true variance beyond this assumption; we tested the sensitivity of *P* (*≥* 1) to this assumption under a Negative Binomial distribution with matched mean and a range of illustrative dispersion parameters (Supplementary Table S2).

An exploratory extension estimating co-national diaspora network risk, an illustrative mechanism by which importation risk could concentrate within specific communities rather than disperse into the general population, is described in Supplementary Appendix B; it relies on additional population-proportional mixing assumptions that are not empirically evaluated here and is not part of the framework’s core results.

All analyses were conducted in R (version 4.5.2) using packages tidyverse, readxl, cowplot, and tidycensus.

### 2.4 Evaluation against surveillance data

To assess whether the model’s expected importation intensities are consistent with independently observed case counts, we compared them with CDC surveillance data for two diseases in years other than the June 2026 target period, namely 2024 annual dengue against MMWR travel-associated case counts [28] and 2022 annual measles against NNDSS imported case counts [29], at the nine states containing a World Cup host city (Table 2). Because no World Cup took place in those years, these comparisons do not involve the event-specific stream and instead test the importation core shared by all three tiers, Eq. (1) with background routing. For each evaluation year we used the actual I-94 arrivals by country of residence (monthly for dengue, annual for measles), routed them to host cities with the same T-100 fractions, combined them with that year’s WHO-reported incidence and the same *ρ_d_* ranges and *D_d_* windows as the main analysis, with *T_d_* equal to the length of the incidence period (one month for dengue and one year for measles, so that *p_d_* = *D_d_/T_d_* is 0.030–0.038 for the annual measles run, compared with 0.36–0.46 for June), and summed the resulting city-level Λ within each state. The 2024 dengue comparison is not fully independent of the main analysis, because June 2024 arrivals and dengue incidence also enter the main model’s inputs. Two checks were applied. First, because reported cases reflect only detected, travel-linked infections while Λ estimates the underlying number of infected arrivals, detected or not, the ratio reported/Λ (the implied detection fraction) should lie between 0 and 1 for a well-calibrated, travel-dominated disease; this bound is necessary but not sufficient, since a model that systematically overestimated Λ at every state would satisfy it trivially without demonstrating any real predictive skill. Second, for dengue, where reported counts vary enough across states to be informative, we tested whether states with higher modeled Λ also had higher reported case counts using Spearman rank correlation; unlike the bounding check, this tests the model’s ability to discriminate between states rather than only its overall scale, and is the stronger of the two pieces of evidence.

**Table 2:** Comparison of modeled importations with state-level CDC surveillance data at the nine states containing a World Cup host city. Λ is the model’s median expected number of imported cases (95% UI) for the evaluation year, computed from ordinary travel without the World Cup schedule (see Methods); the implied detection fraction is reported cases divided by median Λ.

| Disease | State | $\Lambda$ (median, 95% UI) | Reported | Detection fraction |
| --- | --- | --- | --- | --- |
| Dengue (2024) | FL | 8516 (4042–22,718) | 959 | 0.113 |
|  | TX | 5781 (2744–15,422) | 239 | 0.041 |
|  | NY | 4529 (2150–12,083) | 338 | 0.075 |
|  | CA | 2756 (1308–7351) | 702 | 0.255 |
|  | GA | 2448 (1162–6532) | 56 | 0.023 |
|  | MA | 449 (213–1199) | 143 | 0.318 |
|  | WA | 225 (107–602) | 67 | 0.297 |
|  | PA | 141 (66.8–376) | 66 | 0.469 |
|  | MO | 49.1 (23.3–131) | 12 | 0.245 |
| Measles (2022) | NY | 1.40 (1.01–2.11) | 0 | 0 |
|  | CA | 0.642 (0.46–0.97) | 0 | 0 |
|  | GA | 0.306 (0.22–0.46) | 0 | 0 |
|  | TX | 0.019 (0.013–0.028) | 0 | 0 |
|  | MA | 0.016 (0.011–0.024) | 0 | 0 |
|  | FL | 0.014 (0.010–0.021) | 0 | 0 |
|  | WA | 0.0085 (0.0061–0.0128) | 1 | 118 |
|  | PA | 0.0032 (0.0023–0.0048) | 0 | 0 |
| | MO | $1.7 \times 10^{-5}$ ($1.2$–$2.6 \times 10^{-5}$) | 0 | 0 |

## 3 Results

Unless otherwise noted, results below refer to the schedule-driven model (M3), which allocates World Cup fan travel by match schedule and provides a schedule-informed proxy for visitor flows, though it remains an allocation model rather than an observed measurement of them. Figure 3 compares M3 with the 2026 baseline (M1), and Supplementary Figures S1 and S2 show the corresponding M1 and WC-adjusted (M2) heatmaps.

Under the schedule-driven model (M3), the probability of at least one dengue importation exceeded 0.99 at every host city except Kansas City (0.91) (Figure 1; equivalent heatmaps for the Baseline (M1) and WC-adjusted (M2) models are shown in Supplementary Figures S1 and S2). Malaria probability exceeded 0.99 at Atlanta, New York, Miami, San Francisco, Boston, and Philadelphia, was 0.99 at Houston and 0.64–0.74 at Los Angeles and Dallas, and was effectively zero at Kansas City and Seattle. Pertussis probability exceeded 0.99 at 10 of 11 cities and dropped sharply only at Kansas City (0.15). Influenza probability exceeded 0.99 at seven cities, was 0.99 at Seattle, and fell to 0.95 at Boston, 0.81 at Philadelphia, and 0.43 at Kansas City. Measles importation probability was 0.35 at New York and at most 0.12 elsewhere.

**Figure 1:**
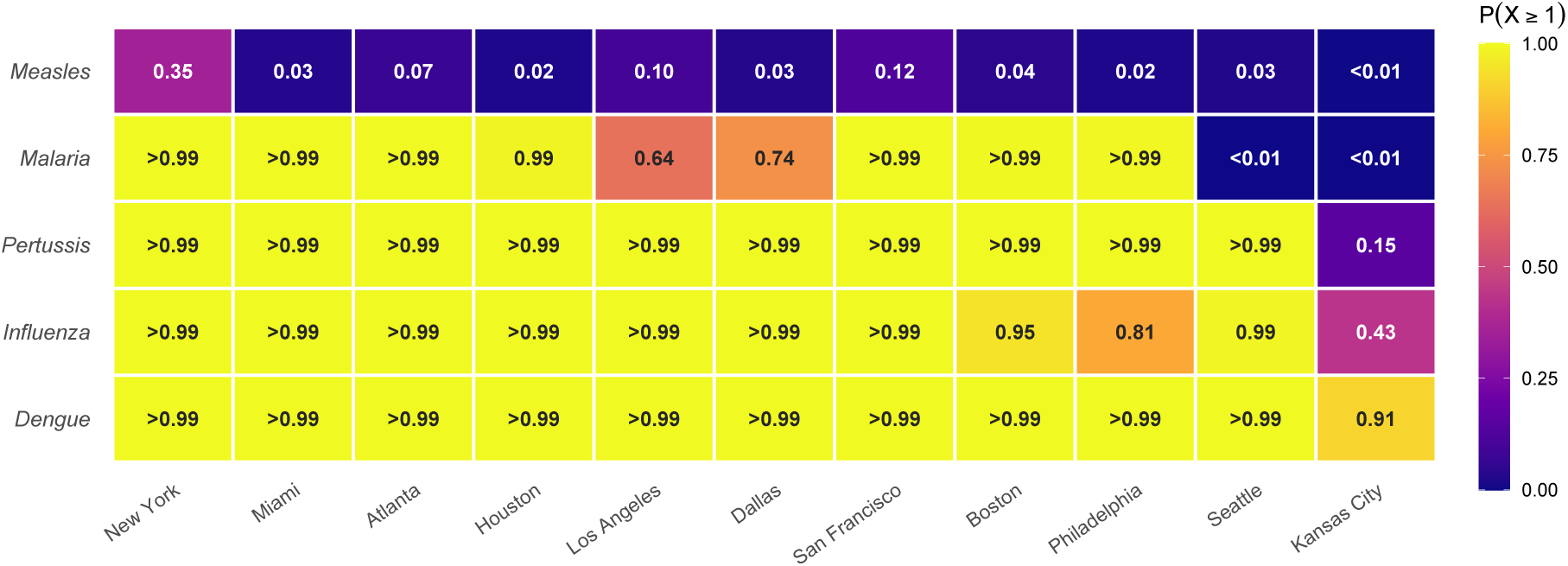
Importation probability for five infectious diseases at 11 US FIFA World Cup 2026 host cities. Cells show the Monte Carlo median of *P* (*≥* 1), the probability of at least one imported case at each city in June 2026, under the schedule-driven model (M3). Cities are ordered by total importation intensity. Values *<* 0.01 and *>* 0.99 are shown as such.

Figure 2 shows Monte Carlo median Λ and 95% uncertainty intervals for each disease at all 11 host cities under the schedule-driven model (M3). For dengue, Miami had the highest absolute importation intensity (Λ = 412, 95% UI 196–1100), followed by New York (274, 95% UI 130–731) and Houston (143, 95% UI 68.1–383); for malaria, Atlanta carried the highest absolute intensity (Λ = 367, 95% UI 113–1007), reflecting Nigeria’s nonstop air service to Atlanta, which carries 62% of Nigeria’s US-bound nonstop passengers in the T-100 data. The 11-city dengue total is about 1.7 times that of malaria and more than three orders of magnitude above that of measles. Interval asymmetry is disease-specific, reflecting the shape of each disease’s *p_d_/ρ_d_* distribution over its parameter ranges (Supplementary Figure S3).

**Figure 2:**
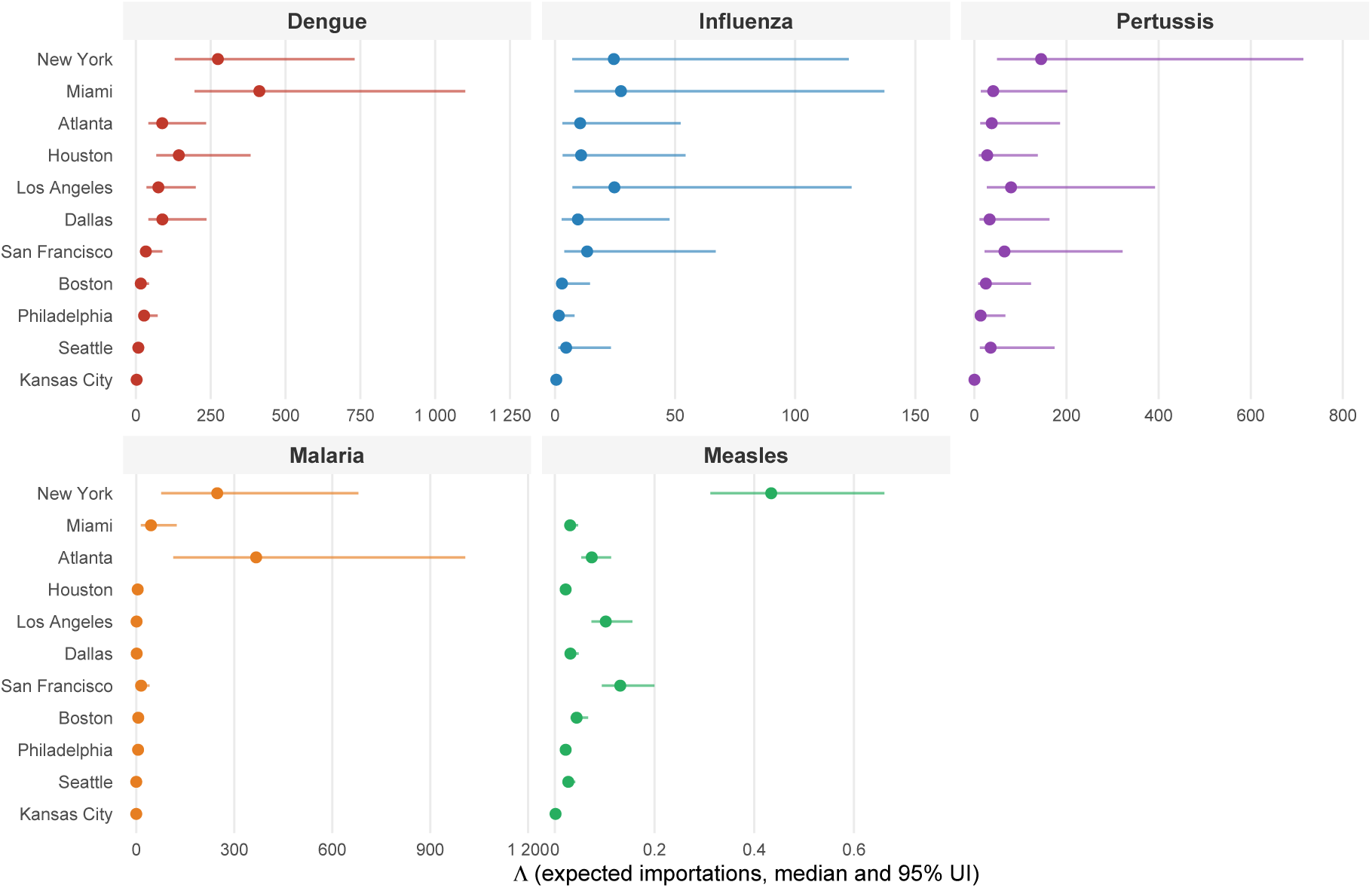
Expected importation intensity (Λ) with 95% Monte Carlo uncertainty intervals by disease, all 11 US host cities, under the schedule-driven model (M3). Points are Monte Carlo medians; bars span the 2.5th–97.5th percentile from 5,000 Uniform draws on *ρ_d_* and *D_d_* (asymmetry examined in Supplementary Figure S3). Each disease panel has an independent x-axis, with cities ordered by overall importation intensity (Figure 1). Corresponding 11-city totals under M1 and M3 appear in Figure 3; per-city heatmaps for M1 and M2 are in Supplementary Figures S1 and S2.

**Figure 3:**
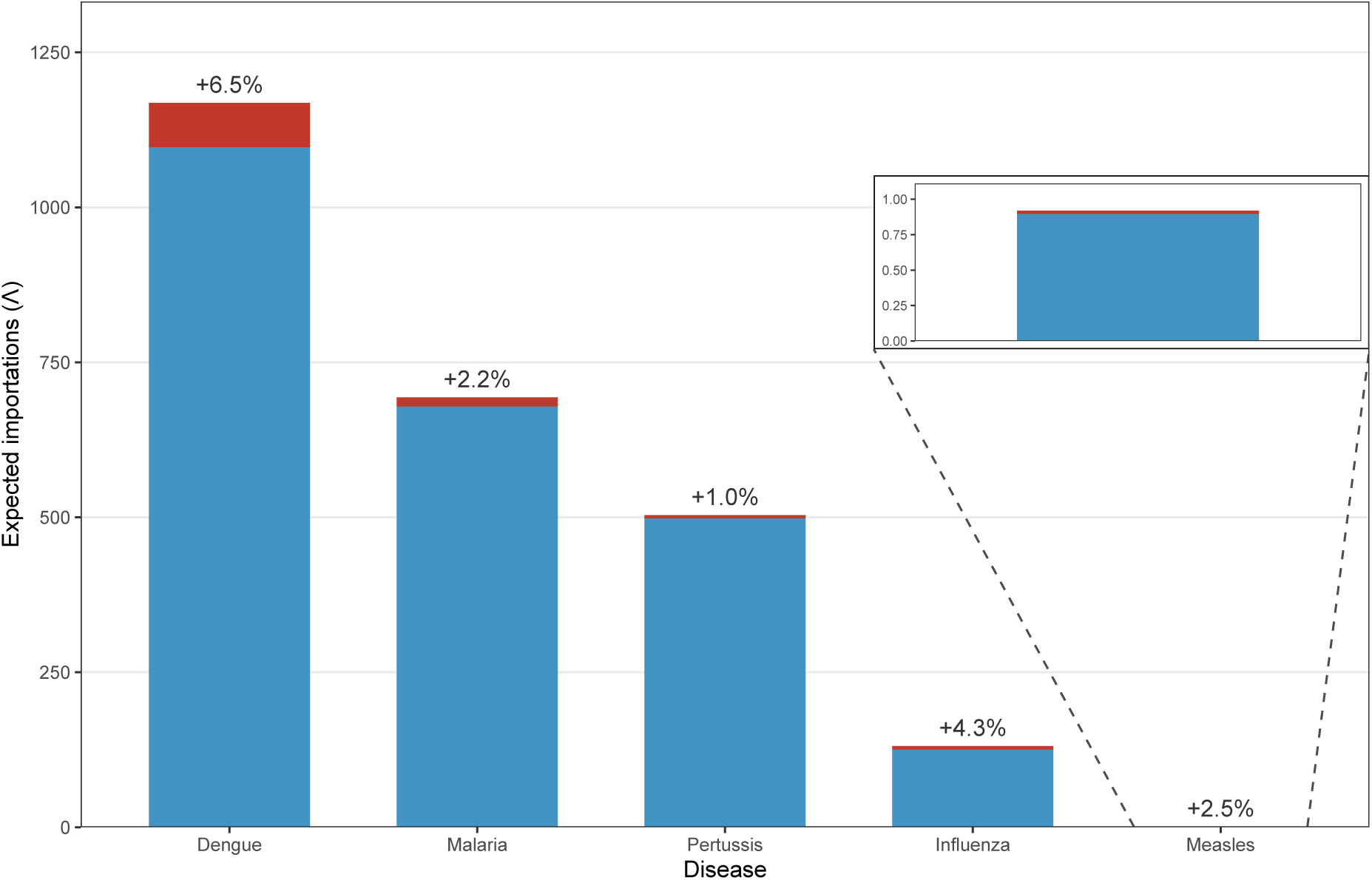
World Cup-specific increment above the 2026 baseline importation risk, by disease. Each bar shows the 11-city Monte Carlo median Λ split into the 2026 baseline without the World Cup (M1, blue) and the schedule-driven WC-specific increment (M3 *−* M1, red). Percentage labels indicate the WC increment as a share of the M1 baseline. The inset shows a zoomed view of the estimates for measles, which are otherwise not visible on the same scale as the other diseases.

Relative to the 2026 baseline without the World Cup (M1 = 1097), the schedule-driven model raised the 11-city dengue total to 1168 (M3, +6.50%), the schedule-informed estimate of the tournament’s event-specific US contribution. Routing the same World Cup-driven growth naively, as if fans distributed across cities like ordinary tourists rather than following the match schedule, gives 1178 instead (M2, +7.41%). Under M2, each country’s World Cup increment reaches the host cities in the same proportion as that country’s ordinary travel, regardless of where its team plays, whereas under M3 it reaches only the US cities hosting that team’s matches, and none of it reaches the United States for matches played in Canada or Mexico (Supplementary Appendix A.7); M3 therefore provides the schedule-informed estimate of the US-specific burden. Comparable patterns held for malaria (M1 = 679 and M3 = 693, +2.15%; naive routing M2 = 699, +2.92%). Across all five diseases, the World Cup’s schedule-driven contribution ranged from 1.0% to 6.5% relative to the 2026-without-WC baseline; in this case study, naive routing overstates this contribution at 2.9% to 8.5%, most for pertussis and least for malaria. Schedule-driven routing also changes where the event’s risk falls. Because M3 sends each qualified team’s fans to the cities where that team plays, importation probability under M3 is well above M2 at several venue cities, for example influenza at Kansas City (0.43 versus 0.05) and malaria at Philadelphia (*>*0.99 versus 0.03) and Boston (*>*0.99 versus 0.63) (Figure 1 and Supplementary Figure S2), even though the 11-city total is lower under M3.

Figure 3 decomposes the 11-city M3 total into the 2026 baseline without the World Cup (M1) and the World Cup-specific increment (M3 *−* M1) for each disease. Malaria and pertussis reached 11-city M3 totals of 693 (95% UI 214–1902) and 503 (95% UI 171–2481), influenza 131 (95% UI 38.6–653), and measles 0.92 (95% UI 0.66–1.40). The measles total, below one expected infected arrival across all 11 cities, reflects the low WHO-reported per-capita incidence across most World Cup source countries; US measles cases arise primarily from outbreak chains seeded by a small number of imports rather than a large number of imported cases. Because M1 already accounts for 94–99% of the M3 total in every disease, most of the absolute importation burden is already present in ordinary 2026 travel, independent of the tournament.

Brazil and Mexico were the dominant dengue contributors overall, and Nigeria and Ghana the dominant malaria contributors (Figure 4). Across all source countries, the World Cup fan stream accounted for about 6% of the 11-city M3 dengue total (Figure 3), so the tournament amplified, rather than created, the importation risk that routine travel from endemic countries already carried. Supplementary Figure S6 ranks source countries by diaspora-weighted importation intensity (Ω*^A^*) rather than raw *λ*; countries with large co-national communities concentrated in their primary destination cities rank higher in Ω*^A^* than in *λ* (for example, Mexico ranks first for dengue by Ω*^A^* but second by *λ*), suggesting where community-level exposure could concentrate under the exploratory assumptions of Supplementary Appendix B.

**Figure 4:**
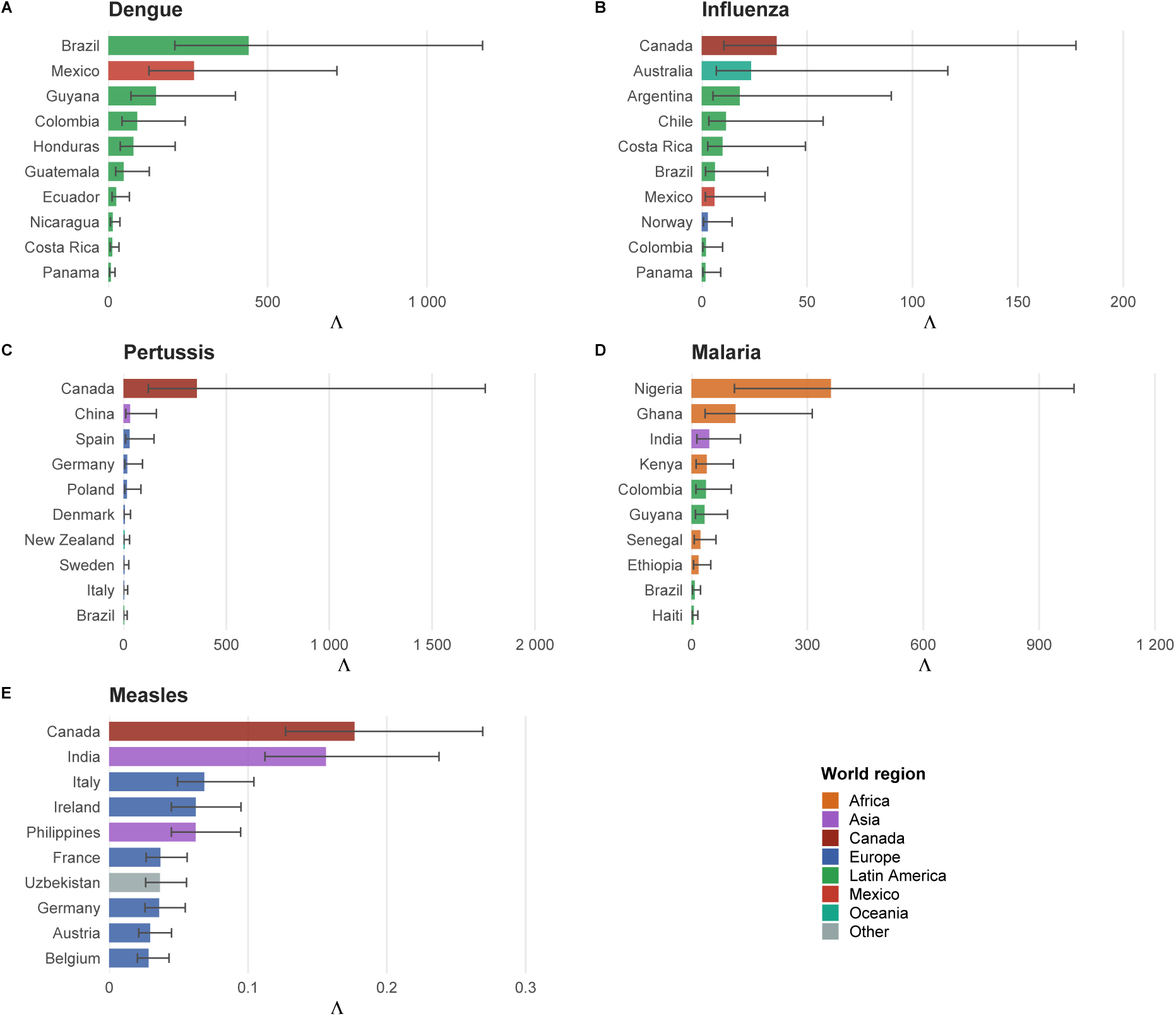
Top 10 source-country contributors to expected importation intensity by disease, under the schedule-driven model (M3). Bars show Monte Carlo median expected importations *λ_c,·,d_* summed across all 11 US venue cities; error bars show 95% Monte Carlo uncertainty intervals. Because *ρ_d_* and *D_d_* are drawn once per disease and shared by all countries, these intervals move together, so the country ranking is more stable than the overlapping bars suggest. Colors denote world region. Panels show (A) dengue, (B) influenza, (C) pertussis, (D) malaria, and (E) measles.

### 3.1 Comparison with surveillance data

Table 2 reports modeled Λ, reported case counts, and the implied detection fraction (reported/Λ) at each of the nine states containing a World Cup host city, for the two diseases and evaluation years described in Methods.

For dengue, modeled Λ and reported cases were significantly rank-correlated across the nine states (Spearman *ρ* = 0.83, *p* = 0.008). Implied detection fractions were also bounded between 0 and 1 at all nine states (2.3–46.9%), consistent with the well-documented under-ascertainment of travel-associated infections in passive surveillance, though this bound alone does not confirm it (see Methods). Georgia was the clearest departure from the otherwise close rank agreement. It ranked fifth by modeled Λ but eighth by reported cases, and had the lowest implied detection fraction of any state (2.3%). This is consistent with T-100’s tendency to over-count arrivals at major connecting hubs such as Atlanta (see Limitations), though a single-state deviation cannot rule out other explanations. For measles, eight of nine states had zero reported imported cases in 2022, and modeled Λ at these states was below 0.7 except at New York (1.40); because reported counts are tied at zero across most states, a rank correlation is not informative here, and this comparison should be read only as a qualitative check that the model does not predict spurious measles risk where none was observed. Washington was the exception, with one reported imported case against a median Λ of 0.0085, an implied detection fraction well above 1, consistent with T-100’s under-counting of arrivals at cities reached mainly via connecting flights (Seattle, in this case). For New York, with Λ = 1.40, a Poisson model gives a probability of exp(*−*1.40) *≈* 25% of observing zero imported cases, so the absence of reported cases there is consistent with chance in a single year.

Pertussis, influenza, and malaria could not be evaluated in the same way. US pertussis case counts are dominated by local transmission chains rather than importation events, so comparison against total reported cases produced implied detection fractions exceeding 100% at multiple states and was not methodologically valid; malaria showed the same failure even more severely, with implied detection fractions exceeding 100% at four states (California, Texas, Massachusetts, and Pennsylvania) and modeled Λ collapsing to zero at two others (Missouri and Washington) that nonetheless reported 23 and 44 cases, consistent with T-100’s connecting-hub routing bias overwhelming the signal at this scale; no comparably structured free surveillance dataset distinguishing imported from locally-acquired influenza cases at state level was identified. These three diseases’ importation estimates should therefore be interpreted with the same caution as the framework’s other untested outputs.

## 4 Discussion

Applied to the WC2026 case study, the framework’s central finding is that the schedule-informed estimate of the tournament’s event-specific US contribution to importation risk, properly isolated using a 2026 baseline without the World Cup (M1) and schedule-driven allocation of fan travel (M3), is modest, 1.0–6.5% of expected importations across the five diseases studied (Figure 3). Because M1 already accounts for 94–99% of the M3 total in every disease, most of the absolute importation burden is present in ordinary 2026 travel regardless of the tournament. Naively routing the same World Cup-driven growth as ordinary background travel (M2), without accounting for which host cities each qualified team’s matches are actually played in, instead suggests a contribution of 2.9–8.5%, from about 1.1 times (dengue) to nearly 9 times (pertussis) the schedule-driven estimate; because M2 spreads fans along ordinary travel routes regardless of where their teams play, it misplaces the event’s contribution among host cities and, in this case study, also overstates its total, in part because the co-hosts Canada and Mexico, both major source countries, played their group matches at home. This gap illustrates why schedule-driven allocation, not just an updated travel-volume projection, is necessary for accurate mass-gathering risk assessment. Year-round monitoring of international travelers from endemic regions therefore remains indispensable regardless of the tournament; targeted tournament-period surveillance is a complement, not a substitute. Within this case study, dengue, malaria, and pertussis emerged as the dominant disease-specific risks, with dengue concentrated in Miami, New York, and Houston, malaria in Atlanta and New York, and pertussis in New York, Los Angeles, and San Francisco.

Dengue was the dominant disease-specific importation threat, driven by high June dengue burden in Latin American source countries, led by Brazil and Mexico, and large bilateral travel volumes from those countries. The 11-city dengue total carries a right-skewed uncertainty interval (median 1168, 95% UI 555–3117), so plausible values extend farther above the median than below it.

In 2024, 3,798 dengue cases were reported in the US, 3,693 (97.2%) of them travel-associated, a 359% increase over the 2010–2023 annual average [28], consistent with the high modeled baseline. Routine bilateral travel from endemic Latin American nations continues to sustain this pre-existing risk. Because the schedule-driven model (M3) routes fan travel to the venues where matches are actually played rather than along ordinary travel routes, preparedness resources calibrated to M3, rather than the WC-adjusted model (M2), are directed toward the cities where event-related fan presence is expected to concentrate.

Standard importation models also treat all arrivals identically regardless of the social environment they enter, whereas an imported case might instead concentrate within a co-national community, facing language or healthcare-access barriers and gathering-driven exposure not captured by city-level *P* (*≥* 1) alone. A supplementary, exploratory extension (Appendix B) suggests this concentration is a plausible additional pathway under a proportional-mixing assumption, but does not model onward transmission explicitly and its assumptions have not been empirically evaluated; we present it as motivation for future work rather than a core finding of this study.

We are not aware of any laboratory-confirmed case of dengue, malaria, measles, pertussis, or influenza publicly attributed to World Cup-related travel by US public health authorities as of October 2026. This absence should not be read as evidence that the estimated risk failed to materialize; surveillance for travel-associated infections depends on a chain of behaviors (a symptomatic traveler seeking medical care, being tested, and having that result linked back to a specific trip), and each step filters out cases. Visitors on short trips may leave before symptoms begin, particularly for dengue and malaria, whose incubation periods (Section 2.2) can exceed a typical spectator’s stay. This ascertainment gap is a structural limitation of passive travel-medicine surveillance generally, not a flaw unique to World Cup-related reporting. Consequently, the framework’s outputs (Λ, *P* (*≥* 1)) are best interpreted as expected *infected-arrival* counts, not as predictions of confirmed US case counts; reconciling the two would require an additional US-side detection-probability correction, distinct from the source-country under-reporting parameter *ρ_d_* used here.

Our analysis has several limitations. First, Poisson assumes independence and equates mean with variance; correlated arrivals from coordinated group travel by fans from the same country introduce overdispersion, so the Poisson model understates the variability of the actual count around Λ. This count variability is distinct from the parameter uncertainty in Λ itself, which is what the reported 95% UIs describe. A Negative Binomial robustness check with matched mean (Supplementary Table S2) shows this matters little for the paper’s dominant findings, since *P* (*≥* 1) at the highest-Λ city for each disease stays above 0.99 for dengue, malaria, and pertussis and at 0.96 for influenza even under substantial illustrative overdispersion (*θ* = 1). It matters more at low-intensity, borderline cities, where *P* (*≥* 1) can fall substantially (for example, dengue at Kansas City from 0.91 to 0.71 at *θ* = 1); estimates at these cities should be read with correspondingly more caution. Second, *ρ_d_* corrects for under-reporting in source-country surveillance but does not model US detection rates; our Λ estimates represent the underlying number of infected arrivals regardless of detection status, not confirmed US case counts, so comparisons with CDC surveillance tallies require additional adjustment. Third, *C_c,d_/P_c_* uses totalpopulation incidence without age stratification, which introduces directional biases that vary by disease; malaria risk from high-endemic sub-Saharan African settings is likely overestimated because pediatric cases dominate reported burden while adult travelers typically carry partial acquired immunity through repeated prior exposure. Fourth, the analysis covers only the 11 US host cities; Canadian and Mexican venues, which receive meaningful World Cup fan flows, are outside the primary scope. Fifth, T-100 records only nonstop international segments, so passengers on connecting itineraries are attributed entirely to their entry airport rather than their true final destination; this likely inflates estimated risk at major connecting hubs (for example, Atlanta) and understates it at cities reached mainly via connecting flights (for example, Seattle and Kansas City). This directional bias is directly visible in the comparison with surveillance data (Table 2), where Georgia’s implied detection fraction is the lowest of any dengue state (over-counted denominator), while Washington’s is the only one exceeding 1 for measles (under-counted denominator). Reconstructing the final destinations of international passengers would require itinerary data that free sources do not provide, since the BTS Airline Origin and Destination Survey (DB1B) covers only domestic itineraries of US carriers; we accept this as a limitation of the routing approach rather than resolve it with commercial data that would compromise the framework’s low-cost, widely reproducible design. Sixth, the measles estimate counts independent seeding events and does not represent the local outbreak chains that actually drive US measles case counts. Seventh, the World Cup growth factors *ϕ_c_* are derived from NTTO point projections and treated as deterministic; uncertainty in fan-travel forecasts is not propagated into the reported intervals, so the true uncertainty bounds are likely wider than reported for M2 and M3 estimates. The World Cup increment is also defined as growth above the background trend for all 12 NTTO markets, including China, India, and Italy, which did not qualify; their above-trend growth is therefore counted under M2 but, with no matches to route it to, not under M1 or M3 (Appendix A.5). In addition, the match schedule used for *ω_c,v_* was the group-stage template available before the six playoff qualifiers (Bosnia and Herzegovina, Czech Republic, DR Congo, Iraq, Sweden, and Turkey) were known, so their fan streams are not routed, and Mexico’s three home matches are absent, which raises the US share of South Africa’s matches from one in three to one in two. Finally, because *p_d_* enters multiplicatively it is a key driver of uncertainty; a one-parameter sensitivity analysis (Supplementary Table S1) shows that malaria and influenza estimates are most sensitive to this parameter (fold-change of 5.0*×* and 4.4*×* across the range of *D_d_*), while measles is least sensitive (1.3*×*). The malaria window also excludes long-term asymptomatic carriage by design (see Methods), so malaria estimates describe infections that can become clinical after arrival.

The framework’s data requirements (routine bilateral travel volumes, publicly reported disease incidence, and an event schedule mapping visitor sub-populations to venues and dates) are freely available for mass gathering events hosted in the United States, where none of these inputs requires a paid data license or specialized modeling infrastructure, and other host countries can apply the framework where equivalent arrival and routing data are published. Applying it to a new event requires three substitutions, namely (i) updated country-of-residence arrival volumes and routing fractions for the host country or cities; (ii) current disease incidence and under-reporting/travel-capable parameter ranges for the pathogens of local concern; and (iii) an event-specific allocation rule analogous to the schedule-driven routing fraction *ω_c,v_* (Appendix A.4), which need not be a sports fixture list, since for religious pilgrimages, trade expositions, or multi-city festivals it reduces to whatever public schedule or ticketing allocation determines where and when international attendees arrive. Because the three model tiers (M1–M3) share identical disease-data inputs and differ only in travel-volume assumptions, the same code base separates any new event’s marginal contribution from its host cities’ pre-existing baseline risk, the same decomposition demonstrated here for WC2026. The Olympic Games, the Hajj, and other recurring mass gatherings with a fixed international attendee schedule and publicly available host-country travel and surveillance data are natural next applications. Because no step in the pipeline depends on proprietary or fee-based data, the framework is particularly well suited to public health agencies operating under budget constraints, including in lowand middle-income host countries that publish such data, where commercial mobility products or dedicated simulation infrastructure may be unaffordable or unavailable ahead of an event.

## 5 Conclusions

We presented a schedule-driven, uncertainty-aware framework for estimating infectious disease importation risk at mass gathering events, and demonstrated it using the 2026 FIFA World Cup group stage as a case study. Because the framework decomposes observed travel into a pre-existing baseline and an event-specific increment using only free, publicly available data, it can be transferred to future mass gathering events for which equivalent travel and surveillance data are available; a public health agency can substitute updated inputs to obtain a first-pass, event-specific risk assessment without specialized modeling infrastructure or a commercial data license.

## Supporting information

Detailed math and definitions

## Data Availability

All data produced are available online at the GitHub link in the manuscript and available freely

https://github.com/jdiestra1977/FIFA_worldCup_2026_risk

## Ethical approval

This study did not involve human subjects, animal subjects, or the collection of primary or identifiable data. All data used were obtained from publicly available, de-identified, aggregate sources (WHO surveillance reports, US Customs and Border Protection I94 statistics, Bureau of Transportation Statistics T-100 data, and US Census Bureau American Community Survey estimates). Ethical approval was therefore not required.

## Data availability

All data used in this study are publicly available. Disease incidence data were obtained from the World Health Organization (Global Dengue Surveillance, World Malaria Report 2024, Immunization Data, and FluNet). Travel data were obtained from US Customs and Border Protection I-94 arrivals, Bureau of Transportation Statistics T-100 records, National Travel and Tourism Office forecasts, and the published FIFA match schedule. National population estimates were obtained from Worldometers, surveillance counts used for evaluation from the US Centers for Disease Control and Prevention (MMWR and NNDSS), and diaspora data from the US Census Bureau American Community Survey 5-year estimates (Table B05006). Analysis code is available at https://github.com/jdiestra1977/FIFA_worldCup_2026_risk.

## CRediT authorship contribution statement

**Jose L. Herrera-Diestra:** Conceptualization, Data curation, Formal analysis, Investigation, Methodology, Software, Supervision, Validation, Visualization, Writing – original draft, Writing – review & editing. **Nyall Jamieson:** Writing – original draft, Writing – review & editing. **Mallory J. Harris:** Visualization, Writing – original draft, Writing – review & editing. **Kaiming Bi:** Writing – original draft, Writing – review & editing. **Martial L. Ndeffo-Mbah:** Writing – original draft, Writing – review & editing. **Melissa Zeynep Ertem:** Writing – review & editing. **Amera Al-Amery:** Writing – review & editing. **Lauren Ancel Meyers:** Supervision, Writing – review & editing.

## Declaration of generative AI and AI-assisted technologies in the manuscript preparation process

During the preparation of this work, the author(s) used Claude (Anthropic) to assist with editing and reformatting portions of the manuscript text. After using this tool, the author(s) reviewed and edited the content as needed and take full responsibility for the content of the published article.

## Acknowledgements

J.L.H-D. acknowledges the support of the Department of Mathematics at Tarleton State University.

