## Supplementary material for "A schedule-driven importation risk framework for mass gathering events: FIFA World Cup 2026": Detailed math and definitions

[1] Department of Mathematics, Tarleton State University, Stephenville, TX 76402, USA
[2] Department of Integrative Biology, University of Texas at Austin, Austin, TX 78712, USA
[3] Department of Biology, University of Maryland, College Park, MD 20742, USA
[4] University of Maryland Institute for Health Computing, North Bethesda, MD 20852, USA
[5] Department of Management, Policy and Community Health, School of Public Health, The University of Texas Health Science Center at Houston, Houston, TX 77030, USA
[6] Department of Veterinary Integrative Biosciences, College of Veterinary Medicine & Biomedical Sciences, Texas A&M University, College Station, TX 77843, USA
[7] Thomas J. Watson College of Engineering, State University of New York at Binghamton, NY, 13850, USA
[8] Department of Computer Information Systems, Jordan University of Science and Technology, Irbid, Jordan

### Appendix A. Mathematical specification of travel flow estimates $N_{c,v}$

#### A.1 Notation

| Symbol | Definition |
|---|---|
| $c$ | Source country ($c = 1, \ldots, C$) |
| $v$ | US venue city ($v = 1, \ldots, 11$) |
| $A_c$ | June 2024 COR arrivals from country $c$ to the US (I-94 program, CBP) |
| $A_{c,2026}^{\text{NTTO}}$ | NTTO-projected June 2026 arrivals from country $c$ (Top-12 inbound markets only; see A.3) |
| $T_{c,v}$ | June passengers from country $c$ to US city $v$, summed over June 2023–2025 (BTS T-100 Form 41 data); $T_c^{\text{US}}$ is the corresponding total to all US airports |
| $f_{c,v}^T$ | T-100 routing fraction: share of country $c$'s US-bound passengers landing at city $v$ (see A.2). Because the 11 cities receive only part of this traffic, $\sum_v f_{c,v}^T \leq 1$. |
| $\phi_c$ | Country-specific 2026 growth factor (see A.3) |
| $g_c$ | Background (non-WC) trend component of $\phi_c$, representing non-WC travel growth from 2024 to 2026 |
| $m_{c,v}$ | Number of group-stage matches that country $c$ plays at US venue $v$ |
| $M_c$ | Total group-stage matches for country $c$ across all WC venues ($M_c = 3$ for all qualified teams) |
| $\omega_{c,v}$ | Schedule-driven routing fraction for WC fans from country $c$ to US venue $v$ (see A.4) |
| $N_{c,v}$ | Projected June 2026 arrivals from country $c$ to venue city $v$ (model-tier specific) |

#### A.2 T-100 routing fraction $f_{c,v}^T$

The T-100 routing fraction gives the proportion of country $c$'s US-bound passengers that land at each venue city $v$:

$$f_{c,v}^T = \frac{T_{c,v}}{T_c^{\text{US}}}$$

where $T_{c,v}$ is the number of June passengers from country $c$ to US city $v$ and $T_c^{\text{US}}$ the number of June passengers from country $c$ to all US airports, both summed over June 2023–2025 (BTS T-100 Form 41 data). Because the 11 venue cities receive only part of each country's US-bound traffic, $\sum_v f_{c,v}^T \leq 1$, and the remainder corresponds to US destinations outside the study. The fractions $f_{c,v}^T$ are fixed wherever background routing is used. M1 and M2 route their respective total travel volumes entirely through $f_{c,v}^T$, whereas M3 routes the background stream through $f_{c,v}^T$ and the event-specific stream through $\omega_{c,v}$ (A.4).

#### A.3 Growth factor $\phi_c$ and its components

The growth factor

$$\phi_c = \frac{V_{c,2026}}{V_{c,2024}}$$

scales June 2024 baseline arrivals to projected June 2026 levels, where $V_{c,t}$ denotes projected total June visitors from country $c$ in year $t$, concretized differently for each of the three country classes below (via $A_c$ and, for Top-12 markets, $A^{\text{NTTO}}_{c,2026}$; see A.1). Three country classes are distinguished.

1. **Top-12 inbound markets** (Australia, Brazil, Canada, China, France, Germany, India, Italy, Japan, Mexico, South Korea, UK), using NTTO country-specific projections:
$$\phi_c = \frac{A^{\text{NTTO}}_{c,2026}}{A_{c,2024}}$$
The World Cup effect is already embedded in the NTTO forecast. The background trend component is the same data-derived factor used for all countries, $g_c = \phi^{\text{bg}} = 1.077$, so the WC increment of a Top-12 market is $\phi_c - 1.077$. This applies to all 12 markets, including China, India, and Italy, which did not qualify (see A.5).

2. **Other WC-qualified nations** use a uniform World Cup growth factor:
$$\phi_c = \phi^{\text{WC}} = \frac{85{,}017}{72{,}390} = 1.174$$
the ratio of NTTO's forecast of total international arrivals to the United States in 2026 (85,017 thousand) to arrivals in 2024 (72,390 thousand), which includes the anticipated World Cup uplift (NTTO 2025 forecast). The background trend component is $g_c = \phi^{\text{bg}} = 1.077$, so the WC-specific fan increment is $\phi_c - g_c = 0.097$.

3. **Non-qualified nations outside the Top-12 markets** receive no WC adjustment:
$$\phi_c = g_c = \phi^{\text{bg}} = 1.077$$
where $\phi^{\text{bg}}$ is the data-driven background growth rate estimated from COR and T-100 data (June 2023–2025). By construction, $\phi_c - g_c = 0$ for non-qualified countries.

##### A.4 Schedule-driven routing fraction $\omega_{c,v}$

For WC-qualified nation $c$, the fan stream is routed proportionally to the share of its group-stage matches played at each US venue:

$$\omega_{c,v} = \frac{m_{c,v}}{M_c} = \frac{m_{c,v}}{3} \quad \text{(US venues only)}$$

If country $c$ plays no matches at US venue $v$ then $\omega_{c,v} = 0$. Matches played at Canadian or Mexican venues contribute to $M_c$ but not to any US $\omega_{c,v}$, so:

$$\sum_{v=1}^{11} \omega_{c,v} = \frac{\text{(US matches of } c)}{3} \leq 1$$

with strict inequality for nations playing one or more matches outside the US. For non-qualified nations no WC fan stream exists and $\omega_{c,v}$ is not used.

##### A.5 Travel decomposition for M3

Model 3 separates total projected 2026 arrivals from country $c$ into a WC fan stream and a background stream:

$$N_c^{\mathrm{WC}} = A_c \cdot \max\bigl(0,\ \phi_c - g_c\bigr) \qquad N_c^{\mathrm{bg}} = A_c \cdot \phi_c - N_c^{\mathrm{WC}}$$

By construction $N_c^{\mathrm{WC}} + N_c^{\mathrm{bg}} = A_c \cdot \phi_c$ exactly, for every country. For the typical case $\phi_c \geq g_c$, this simplifies to $N_c^{\mathrm{bg}} = A_c \cdot g_c$, so the background stream grows at the baseline rate, and the WC increment $\phi_c - g_c$ is captured entirely by $N_c^{\mathrm{WC}}$. In the data used here, all 12 NTTO markets have $\phi_c > g_c$, so the case $\phi_c < g_c$ does not arise. For non-qualified countries outside the NTTO markets, $\phi_c = g_c$ by definition, giving $N_c^{\mathrm{WC}} = 0$ and $N_c^{\mathrm{bg}} = A_c \cdot g_c$. For the three non-qualified NTTO markets (China, India, and Italy), $\phi_c > g_c$, so the decomposition assigns them a WC increment $N_c^{\mathrm{WC}} > 0$; because these countries play no matches, $\omega_{c,v} = 0$ and this increment is routed under M2 but not under M3 (main text, Limitations). For typical WC-qualified nations the fan increment represents approximately $(1.174 - 1.077)/1.174 \approx 8.3\%$ of total projected 2026 arrivals.

##### A.6 Travel flow $N_{c,v}$ by model tier

**Baseline (M1).** Projected June 2026 arrivals from the background (non-World Cup) growth component only, distributed by T-100 routing fractions. This is the same background stream used in M3 (A.6, below), without the World Cup fan stream added, and represents 2026 travel volumes in the absence of the tournament:

$$\boxed{N_{c,v}^{M1} = N_c^{\mathrm{bg}} \cdot f_{c,v}^{T}}$$

which equals $A_c \cdot g_c \cdot f_{c,v}^{T}$ for the typical case $\phi_c \geq g_c$ (A.5). Using this 2026-projected baseline, rather than raw unprojected 2024 arrivals, isolates the World Cup-specific increment (M3 − M1) from ordinary year-over-year travel growth (M2 − M1); a raw 2024 baseline would conflate the two.

**WC-adjusted (M2).** Projected 2026 arrivals (incorporating the full growth factor $\phi_c$) distributed by T-100 routing fractions. All WC fan travel is assigned to US cities regardless of whether matches are played in Canada or Mexico.

$$\boxed{N_{c,v}^{M2} = A_c \cdot \phi_c \cdot f_{c,v}^{T}}$$

**Schedule-driven (M3).** The background stream is T-100 routed; the WC fan stream is routed by the match schedule:

$$\boxed{N_{c,v}^{M3} = \underbrace{N_c^{\mathrm{bg}} \cdot f_{c,v}^{T}}_{\text{background stream}} + \underbrace{N_c^{\mathrm{WC}} \cdot \omega_{c,v}}_{\text{WC fan stream}}}$$

which equals $A_c\bigl[g_c \cdot f_{c,v}^{T} + (\phi_c - g_c) \cdot \omega_{c,v}\bigr]$ for the typical case $\phi_c \geq g_c$. For non-qualified nations $\phi_c = g_c$, so the fan stream vanishes and $N_{c,v}^{M3} = A_c \cdot g_c \cdot f_{c,v}^{T}$.

##### A.7 Comparing M2 and M3 totals across the 11 US host cities

Let $F_c = \sum_v f^T_{c,v} \leq 1$ be the share of country $c$'s US-bound passengers that land at the 11 host cities (A.2), and $S_c = \sum_v \omega_{c,v}$ the share of its group-stage matches played at US venues (A.4). Summing the M2 and M3 flows of A.6 over the 11 cities and using the identity $N_c^{\text{bg}} + N_c^{\text{WC}} = A_c \phi_c$ gives

$$\sum_v N_{c,v}^{M2} - \sum_v N_{c,v}^{M3} = \big(N_c^{\text{bg}} + N_c^{\text{WC}}\big) F_c - \big(N_c^{\text{bg}} F_c + N_c^{\text{WC}} S_c\big) = N_c^{\text{WC}} \big(F_c - S_c\big).$$

The two tiers therefore differ only through the WC fan stream. M2 assigns a larger 11-city total than M3 whenever $F_c > S_c$, which always holds for countries that play all their group-stage matches in Canada or Mexico ($S_c = 0$), including the co-hosts Canada and Mexico, and M3 assigns the larger total when a country plays all its matches in the United States ($S_c = 1$) and $F_c < 1$. Neither tier bounds the other at the level of individual cities, because M3 concentrates each country's fans at the venues where its team plays (compare Figure 1 of the main text with Supplementary Figure S2). Summed over all source countries, the M2 total exceeded the M3 total for all five diseases in this case study. Importation probability heatmaps for M1 and M2 are shown in Supplementary Figures S1 and S2; these complement the M3 heatmap in Figure 1 and the M1 versus M3 comparison in Figure 3 of the main text.

#### Appendix B. Mathematical specification of the diaspora network extension (exploratory)

This appendix describes an exploratory extension to the core framework, not part of its core results. It illustrates a plausible mechanism by which importation risk could concentrate within a specific co-national community rather than disperse into the general population, under a proportional-mixing assumption applied to Census-derived population concentration. It does not model onward person-to-person transmission explicitly, and its assumptions (in particular, that contact probability is proportional to population share) have not been empirically evaluated. Readers should treat the quantities below as a plausibility argument motivating future empirical work, not as validated risk estimates.

##### B.1 Notation

| Symbol | Definition |
|---|---|
| $D_{c,v}$ | Number of US residents born in country $c$ living in metro area $v$ (ACS 5-year estimates 2019–2023, Table B05006) |
| $\mathrm{FB}_v$ | Total foreign-born population of metro area $v$ (row total of Table B05006) |
| $\kappa_{c,v}$ | Diaspora concentration index: share of metro $v$'s foreign-born population born in country $c$ |
| $\Omega^A_{c,v,d}$ | Expected number of infected arrivals from $c$ at city $v$ that enter the co-national network (Mechanism A) |
| $P^A(\geq 1)_{v,d}$ | Probability that at least one infected arrival enters any co-national network in city $v$ |
| $v_{\text{match}}$ | WC venue city where country $c$ plays |
| $v_{\text{home}}$ | Diaspora hub city (may differ from $v_{\text{match}}$) |
| $\omega_{c,v}$ | Schedule-driven routing fraction: share of $c$'s group-stage matches played at US venue $v$ (Appendix A.4); zero if $c$ does not play at $v$ |
| $\Omega^B_{c,v_h,d}$ | Secondary seeding index for hub city $v_h$, summed over all match venues where $c$ plays (Mechanism B) |

##### B.2 Diaspora concentration index $\kappa_{c,v}$

For each source country $c$ and venue metro area $v$:

$$\kappa_{c,v} = \frac{D_{c,v}}{\mathrm{FB}_v}$$

$\kappa_{c,v} \in [0, 1]$ measures how concentrated the country-$c$ community is *within* the immigrant population of city $v$, the relevant social network for co-national mixing. Summed over all countries of birth, $\sum_c \kappa_{c,v} = 1$ for each $v$; because the model includes only the source countries relevant to the five diseases, the $\kappa_{c,v}$ for the modeled countries sum to less than one in each city.

##### B.3 Mechanism A: local social mixing

Let $X_{c,v,d} \sim \mathrm{Poisson}(\lambda_{c,v,d})$ be the number of infected arrivals from country $c$ at city $v$. Under the proportional-mixing assumption, each arrival independently enters the co-national network with probability $\kappa_{c,v}$. By the Poisson thinning theorem [1], the thinned count is:

$$Y_{c,v,d} \sim \text{Poisson}(\lambda_{c,v,d} \cdot \kappa_{c,v})$$

We define the community-weighted importation intensity:

$$\Omega^A_{c,v,d} = \lambda_{c,v,d} \cdot \kappa_{c,v} = \lambda_{c,v,d} \cdot \frac{D_{c,v}}{\text{FB}_v}$$

Because $Y_{c,v,d}$ values are independent Poisson random variables, the city-level aggregate $\Omega^A_{v,d} = \sum_c \Omega^A_{c,v,d}$ is also a valid Poisson rate. The probability that at least one infected arrival enters any co-national network in city $v$ is:

$$P^A(\geq 1)_{v,d} = 1 - \exp(-\Omega^A_{v,d})$$

This is directly comparable to the main-model probability $P(\geq 1)_{v,d} = 1 - \exp(-\Lambda_{v,d})$. Since $\kappa_{c,v} \leq 1$ for all $c, v$, it follows that $\Omega^A_{v,d} \leq \Lambda_{v,d}$ and therefore $P^A(\geq 1)_{v,d} \leq P(\geq 1)_{v,d}$ always. The diaspora-network probability is a lower bound on the arrival probability, never exceeding it.

##### B.4 Mechanism B: diaspora return seeding

US-resident diaspora members from country $c$ living in hub city $v_{\text{home}}$ may travel domestically to $v_{\text{match}}$ (where country $c$ plays a US match), be exposed to infected co-nationals within the local network that Mechanism A already quantifies, and seed transmission upon return. The secondary seeding index reuses the schedule-driven routing fraction $\omega_{c,v}$ (Appendix A.4) to restrict this pathway to venues where $c$ actually plays, and the local exposure hazard $\Omega^A_{c,v,d}$ (Section B.3) rather than the raw arrival rate $\lambda_{c,v,d}$:

$$\Omega^B_{c,\, v_{\text{home}},\, d} = \sum_{v_{\text{match}} \neq v_{\text{home}}} \omega_{c,\, v_{\text{match}}} \cdot \Omega^A_{c,\, v_{\text{match}},\, d} \cdot \kappa_{c,\, v_{\text{home}}}$$

Because $\omega_{c,v} \leq 1$ and $\kappa_{c,v} \leq 1$ for all $c, v$, and $\Omega^A_{c,v,d}$ is already a well-scaled Poisson rate (Section B.3), $\Omega^B$ remains on the same scale as $\Omega^A$ rather than growing unboundedly, unlike a specification that instead scales a raw diaspora headcount by an assumed match-attendance probability. Summing over source countries gives the hub-city aggregate:

$$\Omega^B_{v_{\text{home}},\, d} = \sum_c \Omega^B_{c,\, v_{\text{home}},\, d}$$

The index $\Omega^B$ is used here only as a relative index of secondary seeding potential. It is not converted into a probability, because the model does not represent attendance, exposure, infection, or onward transmission explicitly.

Because $\omega_{c,v_{\text{match}}} = 0$ whenever country $c$ does not play a match at $v_{\text{match}}$ (Appendix A.4), $\Omega^B$ is defined only for the subset of source countries that are both WC-qualified and scheduled to play at a US venue, 13 of the source countries modeled here meet this condition. A country with substantial $\Omega^A_{c,v,d}$ driven by routine background travel to a city where it is not playing contributes zero to $\Omega^B$ at that venue, since Mechanism B specifically models exposure acquired while attending a match, not general travel; that country's local risk is still fully captured by Mechanism A. This is a deliberate scope restriction consistent with the mechanism's premise, not an omission.

The match venue $v_{\text{match}}$ ranges over the 11 host cities, while the hub city $v_{\text{home}}$ ranges over both the 11 host cities and 15 major non-host metropolitan areas with large immigrant populations (drawn from the same ACS B05006 country-of-birth data), so that secondary seeding into non-venue cities is captured. The 15 non-host hub metropolitan areas are Austin, Charlotte, Chicago, Denver, Detroit, Las Vegas, Minneapolis, Orlando, Phoenix, Portland, Sacramento, San Antonio, San Diego, Tampa, and Washington, DC.

##### B.5 Illustrative results for dengue and malaria

Figure S9 summarizes both mechanisms for dengue and malaria, and Figures S5 and S7 show all five diseases. Under Mechanism A, the probability that at least one imported dengue case enters a co-national diaspora network exceeded 0.99 at Miami, Houston, Los Angeles, and Dallas, was 0.99 at Atlanta and San Francisco and 0.97 at New York, and ranged from 0.38 to 0.59 at the four lowest-intensity cities (Boston 0.59, Seattle 0.59, Philadelphia 0.53, Kansas City 0.38). Malaria was far more concentrated. The probability exceeded 0.99 only at Atlanta, reflecting the city's high malaria importation intensity through Hartsfield-Jackson airport, remained substantial at New York (0.96), Miami (0.92), and San Francisco (0.92), and was below 0.07 at every other host city.

Under Mechanism B, the highest-ranked dengue hubs were the non-host cities Orlando ($\Omega^B = 0.521$) and Tampa (0.294), followed by the host city Boston (0.281), the non-host city Charlotte (0.179), and the host cities New York (0.176), Atlanta (0.157), Houston (0.112), and Philadelphia (0.098). The malaria hub ranking was broadly similar, with values roughly 3- to 6-fold lower depending on the hub (Orlando $\Omega^B = 0.162$, Tampa 0.104, New York 0.064). New York ranked third for malaria rather than fifth, ahead of Boston, reflecting a different balance of qualified source countries between the two diseases. Among source countries that are both WC-qualified and scheduled to play at a US venue, Colombia and Brazil account for nearly all of $\Omega^B$ at every top hub for both diseases (Figure S8), and non-host hubs such as Orlando, Tampa, and Charlotte rank alongside host cities because of their large Colombian and Brazilian diaspora communities. Countries with much larger raw arrival intensities, such as Nigeria for malaria (Figure 4 of the main text), contribute little or nothing to $\Omega^B$ because their risk arrives through routine travel rather than match attendance (Nigeria did not qualify), a pathway that Mechanism A captures but Mechanism B, by design, does not.

These results illustrate a plausible pattern under the extension's assumptions. They have not been empirically validated and should not be read as confirmed community-level risk estimates.

#### Table S1. Sensitivity of expected importations to the travel-while-infected probability $p_d$.

Because $p_d$ enters the model multiplicatively ($\lambda_{c,v,d} = N_{c,v} \cdot C_{c,d}/(P_c \cdot \rho_d) \cdot p_d$), fixing $\rho_d$ at its literature midpoint and varying $p_d$ produces importation totals that scale exactly as $p_d/p_{\text{mid}}$. The table below reports the resulting 11-city totals at $p_{\text{min}}$, $p_{\text{mid}}$, and $p_{\text{max}}$ and the exact fold-change $p_{\text{max}}/p_{\text{min}}$, which equals $D_{\text{max}}/D_{\text{min}}$ because $p_d = D_d/T_d$, for all five modeled diseases.

| **Disease** | $\boldsymbol{p_{\text{min}}}$ | $\boldsymbol{p_{\text{mid}}}$ | $\boldsymbol{p_{\text{max}}}$ | $\boldsymbol{\Lambda(p_{\text{min}})}$ | $\boldsymbol{\Lambda(p_{\text{mid}})}$ | $\boldsymbol{\Lambda(p_{\text{max}})}$ | **Fold** |
|---|---|---|---|---|---|---|---|
| Dengue | 0.197 | 0.312 | 0.427 | 732.79 | 1,160.26 | 1,587.72 | 2.2× |
| Malaria | 0.197 | 0.591 | 0.986 | 230.05 | 690.16 | 1,150.26 | 5.0× |
| Measles | 0.361 | 0.411 | 0.460 | 0.81 | 0.93 | 1.04 | 1.3× |
| Pertussis | 0.230 | 0.509 | 0.789 | 227.19 | 503.07 | 778.95 | 3.4× |
| Influenza | 0.040 | 0.109 | 0.177 | 48.94 | 132.83 | 216.73 | 4.4× |

$\Lambda$ = 11-city total expected importations (M3 schedule-driven model), computed deterministically with $\rho_d$ fixed at its literature midpoint $(\rho_{\text{min}} + \rho_{\text{max}})/2$. Fold $= \Lambda(p_{\text{max}})/\Lambda(p_{\text{min}}) = p_{\text{max}}/p_{\text{min}}$ (exact). Malaria and influenza carry the largest sensitivity to $p_d$; measles the smallest.

#### Table S2. Sensitivity of importation probability to overdispersion.

The main-text Poisson model assumes independent arrivals, so its variance equals its mean; correlated arrivals from fans traveling in groups from the same country would inflate the true variance beyond this assumption (main text Limitations). To test how much this matters, we recomputed $P(\geq 1)$ under a Negative Binomial distribution with the same mean $\Lambda$ but additional dispersion parameter $\theta$ (variance $= \Lambda + \Lambda^2/\theta$; $\theta \to \infty$ recovers the Poisson case), for the highest- and lowest-$\Lambda$ city of each disease for which the Poisson estimate exceeded 0.4. $\theta$ is not fitted to data; the range below is an illustrative bound on plausible group-travel clustering, not an estimated value.

| **Disease** | **City** | $\boldsymbol{\Lambda}$ | $\boldsymbol{P(\geq 1)}$ | | | |
|---|---|---|---|---|---|---|
| | | | **Poisson** | $\boldsymbol{\theta = 5}$ | $\boldsymbol{\theta = 2}$ | $\boldsymbol{\theta = 1}$ |
| Dengue | Miami | 412 | 1.000 | 1.000 | 1.000 | 0.998 |
| Dengue | Kansas City | 2.42 | 0.911 | 0.861 | 0.796 | 0.708 |
| Malaria | Atlanta | 367 | 1.000 | 1.000 | 1.000 | 0.997 |
| Malaria | Los Angeles | 1.02 | 0.639 | 0.605 | 0.561 | 0.505 |
| Pertussis | New York | 145 | 1.000 | 1.000 | 1.000 | 0.993 |
| Pertussis | Philadelphia | 13.6 | 1.000 | 0.999 | 0.984 | 0.932 |
| Influenza | Miami | 27.5 | 1.000 | 1.000 | 0.995 | 0.965 |
| Influenza | Kansas City | 0.561 | 0.429 | 0.412 | 0.390 | 0.359 |

At the highest-$\Lambda$ city for each disease, $P(\geq 1)$ stays above 0.99 for dengue, malaria, and pertussis and at 0.965 for influenza even under substantial overdispersion ($\theta = 1$), so the main text's near-certain importation estimates at major host cities are not an artifact of the Poisson independence assumption. At the lowest-$\Lambda$ cities with a non-trivial Poisson probability, by contrast, $P(\geq 1)$ is materially lower under overdispersion (for example, dengue at Kansas City falls from 0.911 to 0.708 at $\theta = 1$); estimates at these borderline, low-intensity cities should therefore be read with more caution than the paper's dominant, high-confidence findings.

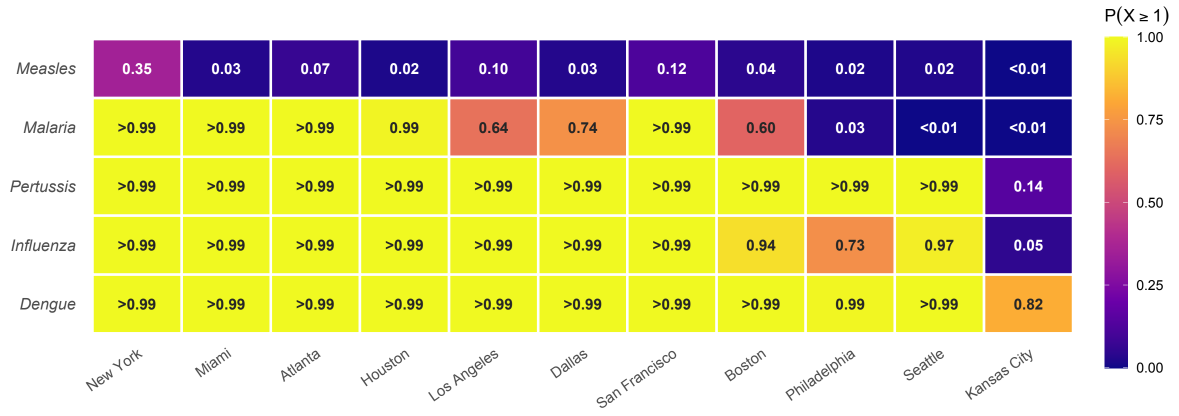


**Figure S1. Importation probability under the Baseline model (M1), 2026 projected travel without the World Cup.** Same layout as Figure 1 in the main text. M1 represents projected June 2026 travel volumes absent the tournament, so that comparison with the schedule-driven model (M3, Figure 1) isolates the World Cup-specific increment from ordinary year-over-year travel growth.

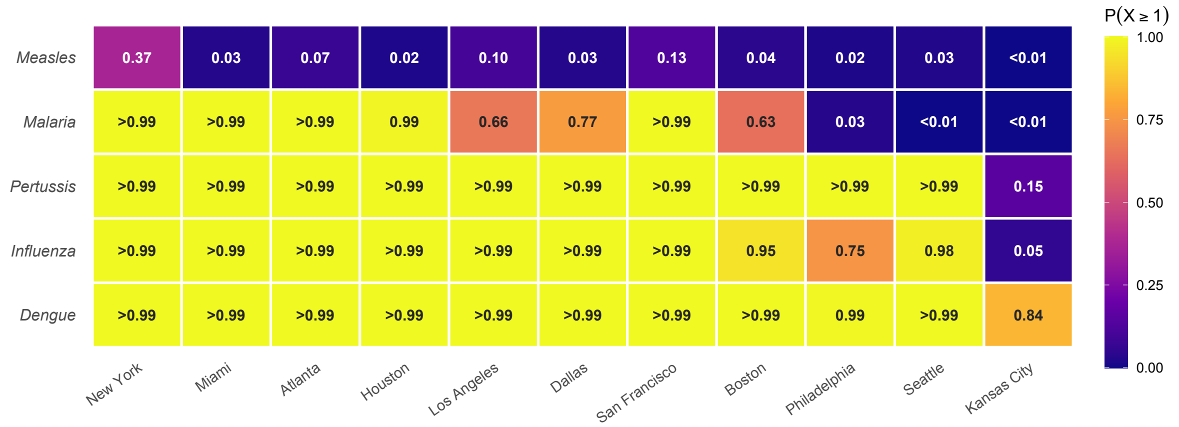


**Figure S2. Importation probability under the WC-adjusted model (M2), June 2026.** Same layout as Figure 1 in the main text. M2 applies World Cup growth factors to national arrivals but distributes all travel to US cities via T-100 routing fractions, including the fan increment for teams playing in Canada and Mexico. In this case study its 11-city total exceeds that of M3 for every disease (Appendix A.7), although at individual venue cities M2 can fall well below M3 because it does not send fans to the cities where their teams play.

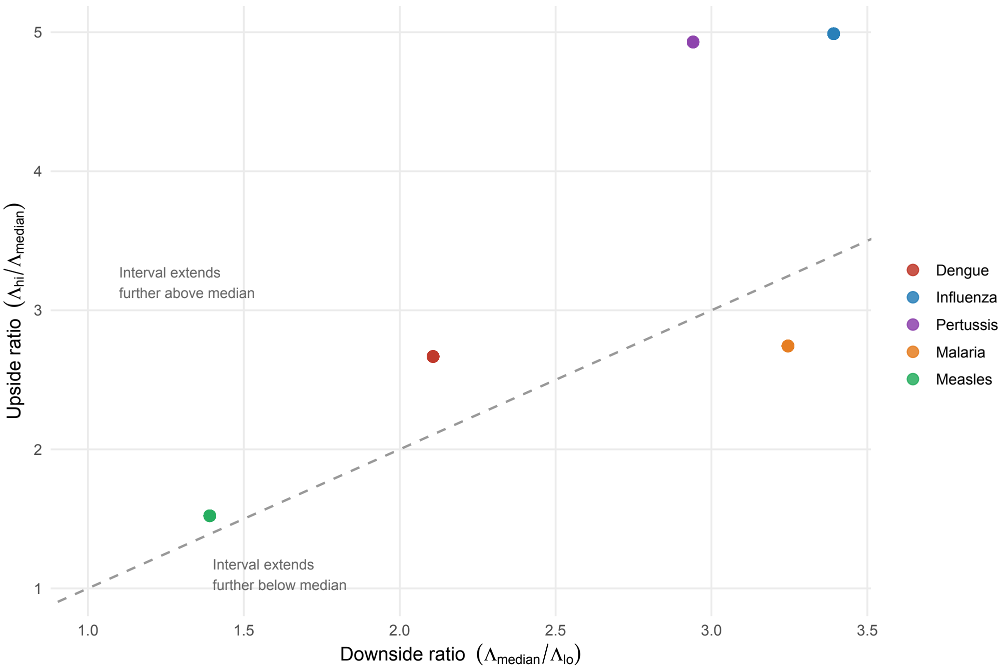


**Figure S3. Direction of parameter uncertainty by disease.** Each point represents one disease's 11-city aggregate. The horizontal axis shows the downside ratio ($\Lambda_{median}/\Lambda_{lo}$), measuring how far the lower end of the 95% UI lies below the median. The vertical axis shows the upside ratio ($\Lambda_{hi}/\Lambda_{median}$), measuring how far the upper end of the 95% UI lies above the median. Points above the dashed diagonal have intervals that extend farther above the median than below it, and points below the diagonal show the reverse.

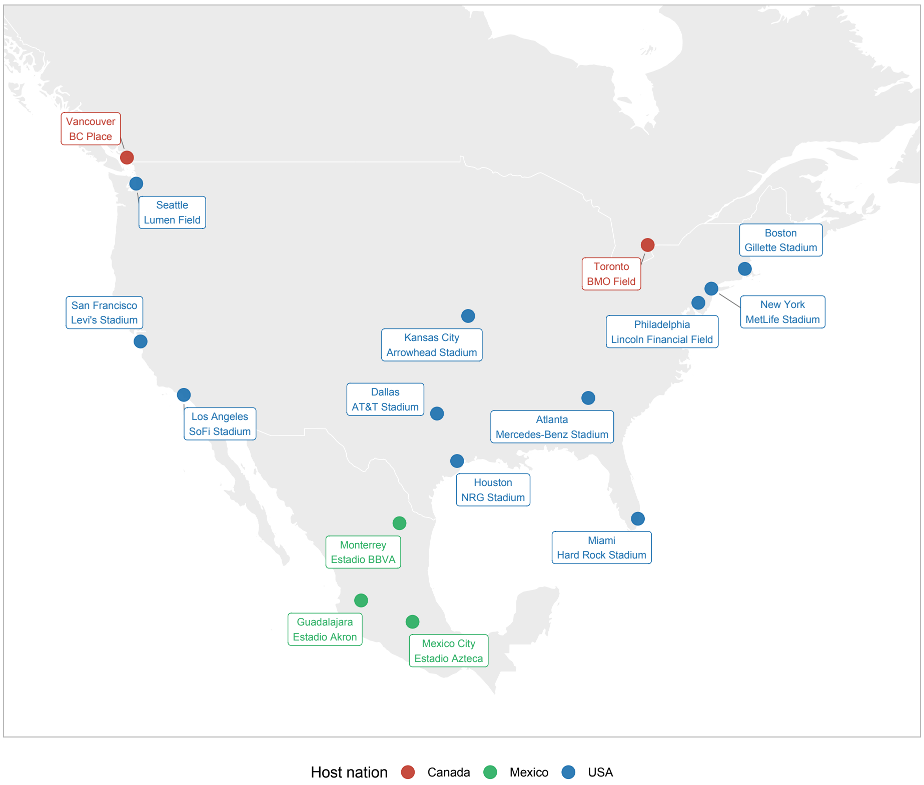


**Figure S4. Geographic distribution of the 16 FIFA World Cup 2026 stadiums across the United States, Canada, and Mexico.** Colors indicate host nation. This analysis focuses on the 11 US venues (blue); Canadian (Toronto, Vancouver) and Mexican (Mexico City, Monterrey, Guadalajara) venues are shown for context. The East Coast cluster (New York, Philadelphia, Boston) is the densest; Kansas City and Seattle are the most geographically isolated US venues. Map lines delineate study areas and do not necessarily depict accepted national boundaries.

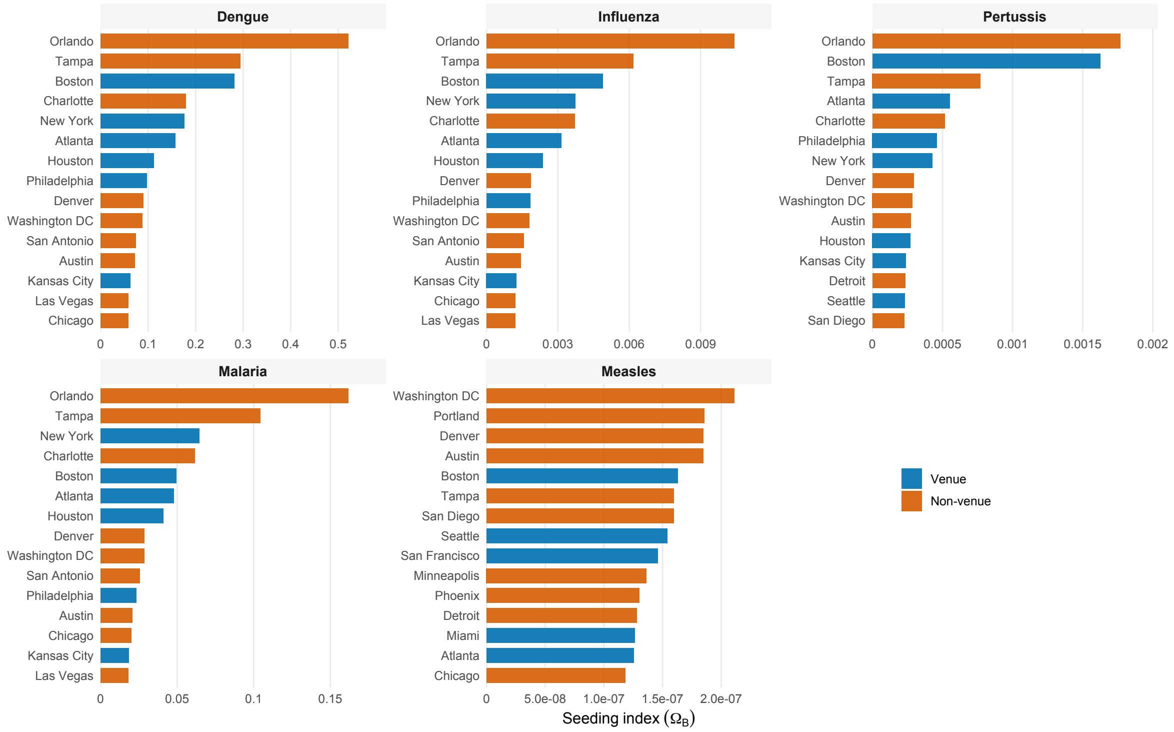


**Figure S5. Secondary seeding risk index ($\Omega^B$) for diaspora hub cities, by disease (Mechanism B).** Bars show the top 15 hub metropolitan areas per disease, ranked by $\Omega^B$ summed across all source countries and match venues, and colored by whether the hub is a World Cup host city or a non-host city. Hubs are the home cities to which diaspora members return after attending matches. $\Omega^B$ is defined only for source countries that are both WC-qualified and scheduled to play at a US venue (Appendix B.4).

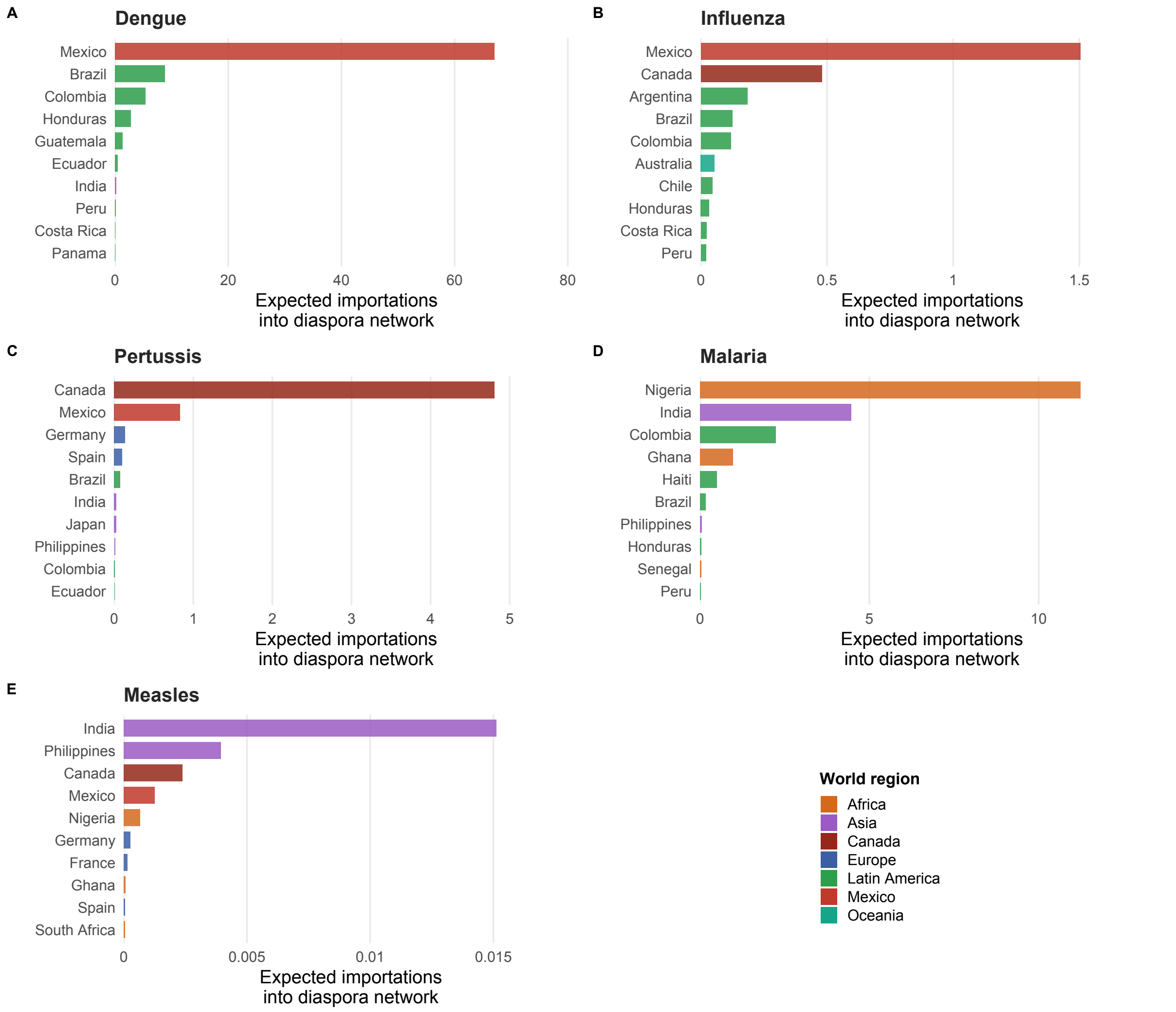


**Figure S6. Top 10 source-country contributors to diaspora-network importation intensity ($\Omega^A$) per disease.** Bars show $\Omega^A$ for each source country, summed across the 11 host cities; colors denote world region. The corresponding raw-$\lambda$ ranking is shown in Figure 4 of the main text.

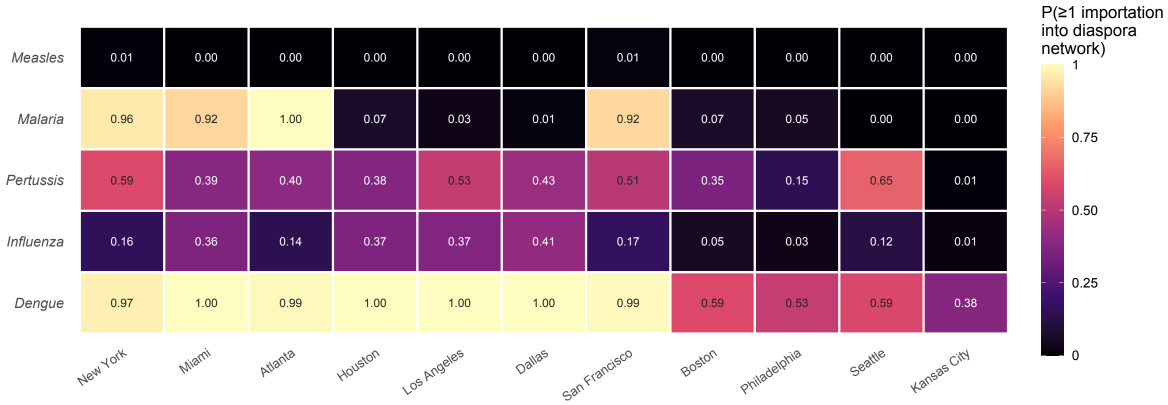


**Figure S7. Diaspora network importation probability ($P^A(\geq 1)$) for all five diseases at 11 US host cities.** Cells show $P^A(\geq 1)_{v,d}$, the probability that at least one imported case enters a co-national diaspora network at each host city. The dengue and malaria rows correspond to Panel A of Figure S9 below.

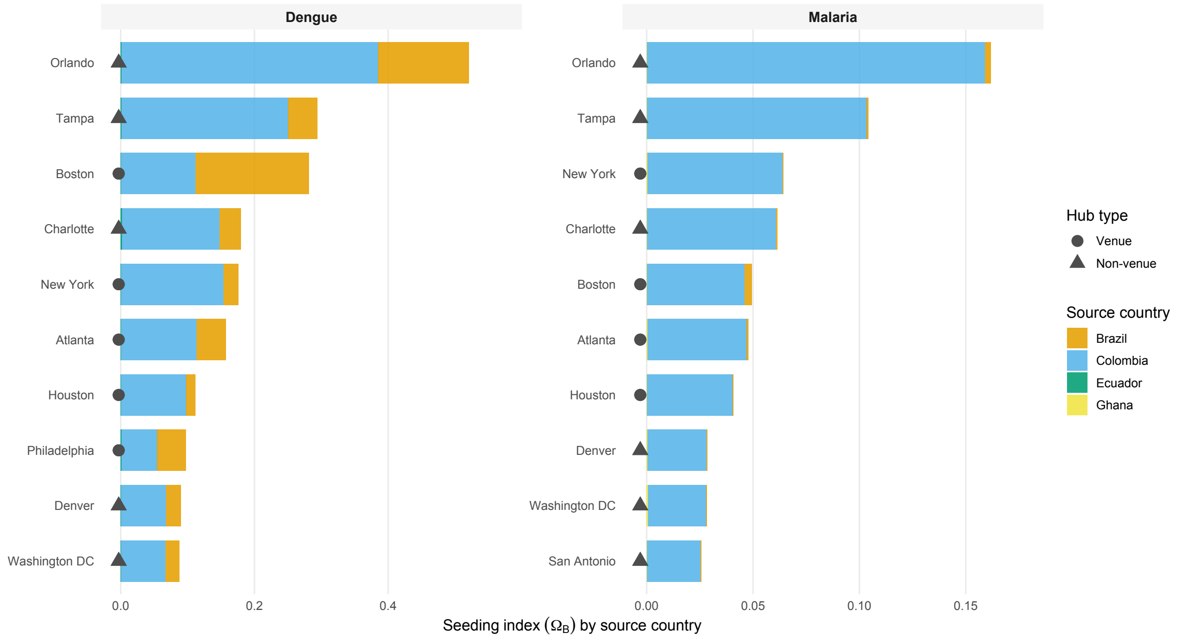


**Figure S8. Source-country contributions to Mechanism B secondary seeding index ($\Omega^B$) for dengue and malaria.** Bars show the source-country contributions to $\Omega^B$ at each of the top hub cities per disease, ranked by total $\Omega^B$. Dot shape distinguishes World Cup venue cities (circle) from non-venue metropolitan areas (triangle). Results are described in Appendix B.5.

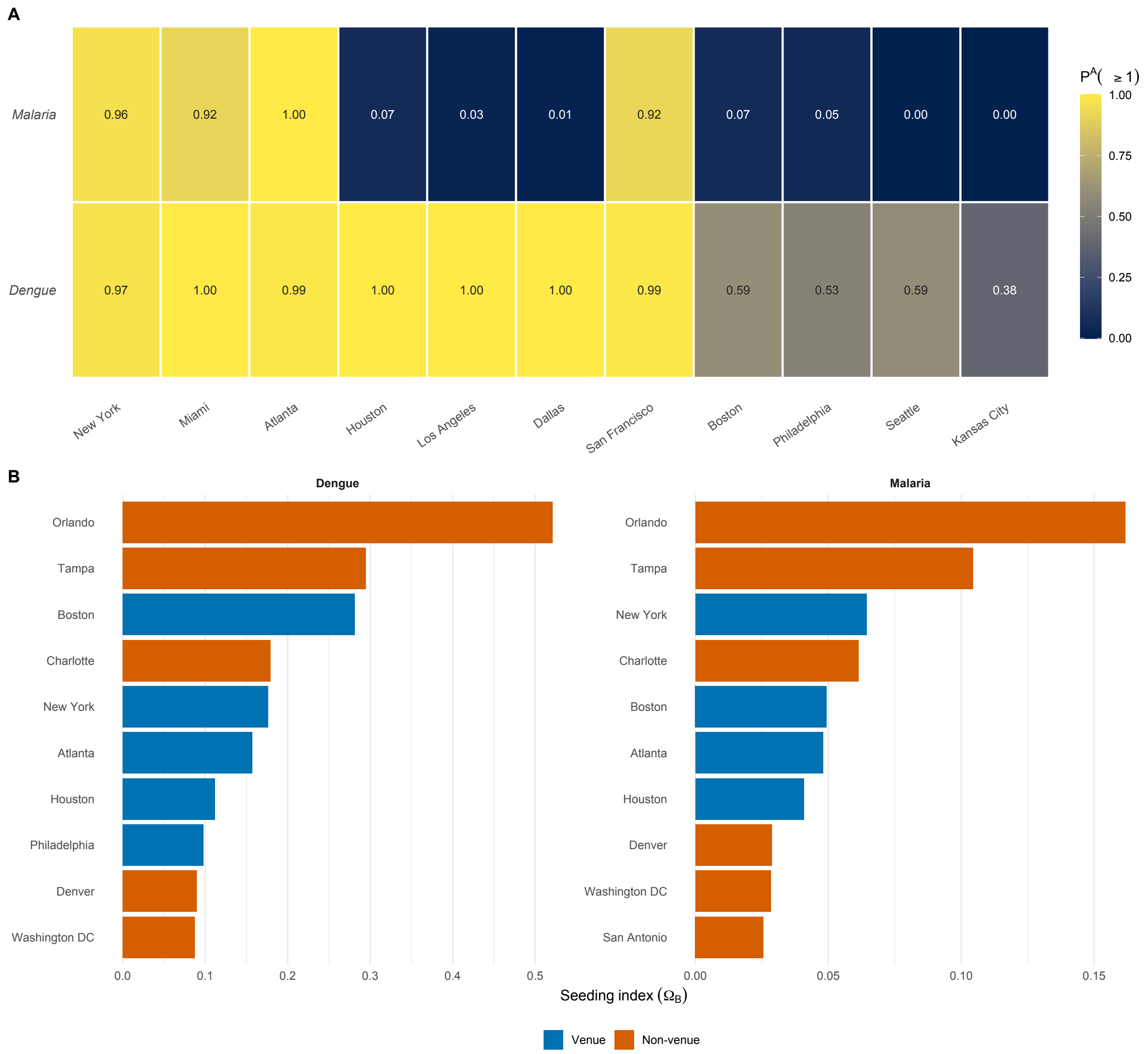


**Figure S9. Exploratory diaspora network seeding risk for dengue and malaria.** (A) Probability that at least one imported case enters a co-national diaspora network at each host city, $P^A(\geq 1)_{v,d}$ (Mechanism A). (B) Secondary seeding index $\Omega^B$ for the top 10 hub metropolitan areas per disease, colored by whether the hub is a World Cup host city (venue) or not (non-venue) (Mechanism B). Results are described in Appendix B.5.
